# Simulation and temporal validation of ICU trajectories after stroke: patient condition, transfer readiness and actual ICU exit

**DOI:** 10.64898/2026.11.17.26368348

**Authors:** Ping Lei, Yanyi Xu, Yuxia Zhang, Nicolas Ray, Ying Gao

## Abstract

**Objective:** To describe how intensive care unit (ICU) time is composed after stroke, and to test whether simulating patient condition, transfer readiness and ICU exit separately predicts as accurately as direct models.

**Materials and Methods:** In 6,366 ICU stays for stroke in MIMIC-IV, hourly readiness criteria divided ICU hours into organ-support needs, stroke-specific needs, and time not explained by these needs. An hourly simulator was built on stays up to 2016, frozen, and validated on 2017–2019 (n=1,714) and 2020 onwards (n=1,521). The primary measure was the 24-hour alive-exit Brier score versus the strongest direct model (gradient boosting).

**Results:** Unexplained time was 34.1% of hours in standard ICUs and 87.6% in neuro intermediate units. 37.8% of stays lost readiness for ≥6 consecutive hours after first confirmation. The simulator and gradient boosting were indistinguishable (Brier 0.1238 vs 0.1245; difference −0.0007, 95% CI −0.0030 to 0.0016). The simulator was better calibrated at 48 and 72 hours but discriminated less well (C statistic 0.71 vs 0.74–0.75). All models overpredicted exit after 2016; the error lay mainly after readiness (median 40 hours simulated vs 68 observed).

**Discussion:** Readiness was not a single event: patients often lost and regained it, and many stayed long after meeting it, especially in neuro intermediate units, which exist for monitoring. Separate modelling did not improve accuracy but placed the post-2016 error after readiness, as the unit mix changed.

**Conclusion:** The simulator matched the strongest direct model while also generating complete trajectories. It needs external validation before any clinical use.

## BACKGROUND AND SIGNIFICANCE

ICU beds are scarce, and patients with acute stroke vary widely in how long they stay. Part of this time reflects medical need: ventilation, vasopressors, control of blood pressure, and monitoring after reperfusion therapy. Part may reflect how care is organised: when decisions are made, where a bed is available, and which units exist in a hospital. Routine data rarely separate the two. This matters both for clinicians deciding when a patient can leave and for informaticians predicting ICU length of stay.

Two gaps motivate this study. First, ICU discharge criteria are mostly static. The Society of Critical Care Medicine guideline defines discharge readiness as no longer meeting ICU admission criteria while meeting the criteria for a lower level of care.[1] Scores such as the Stability and Workload Index for Transfer (SWIFT) assess readiness at one time point.[2] Little is known about when patients first meet such criteria hour by hour, how often they lose them again, and how long they stay after meeting them.

Second, most ICU length-of-stay models predict a single number or a single probability.[3,4] They do not separate the patient’s condition from the decisions of the care team. They cannot produce the path a patient might take, and their errors are hard to trace when care practice changes. Discrete-event simulation models in hospital operations do generate patient paths, but they mostly target operational measures such as waiting times and throughput rather than individual clinical prediction, and fewer than 10% report implementation.[5]

A simulator with separate, observable layers for patient condition, transfer readiness and care decisions could address both gaps.

## OBJECTIVE

In ICU stays for stroke at one centre, we aimed (1) to describe how ICU time divides into time explained by observed clinical needs and time not explained by them, and how transfer readiness evolves hour by hour; (2) to build an hourly simulator of ICU paths with separate layers for condition, readiness and care decisions; and (3) to compare it, in temporal validation, with direct prediction models, and to test the contribution of each layer.

## MATERIALS AND METHODS

### Data and population

We used MIMIC-IV version 3.1 and MIMIC-IV-ED version 2.2, which contain de-identified records from Beth Israel Deaconess Medical Center, Boston, from 2008 to 2022.[6–9] The cohort was the stroke ICU cohort of our earlier study of clinical states (6,368 ICU stays with ischaemic stroke, intracerebral haemorrhage or subarachnoid haemorrhage).[10] We excluded 2 stays in which death was recorded at or before ICU admission, leaving 6,366 stays and 847,383 patient-hours (Table 1). ICU time included time in an explicit list of ICU and neuro intermediate or step-down units (Supplementary Table S1). Stays were followed for up to 30 days (720 hours).

**Table 1.**
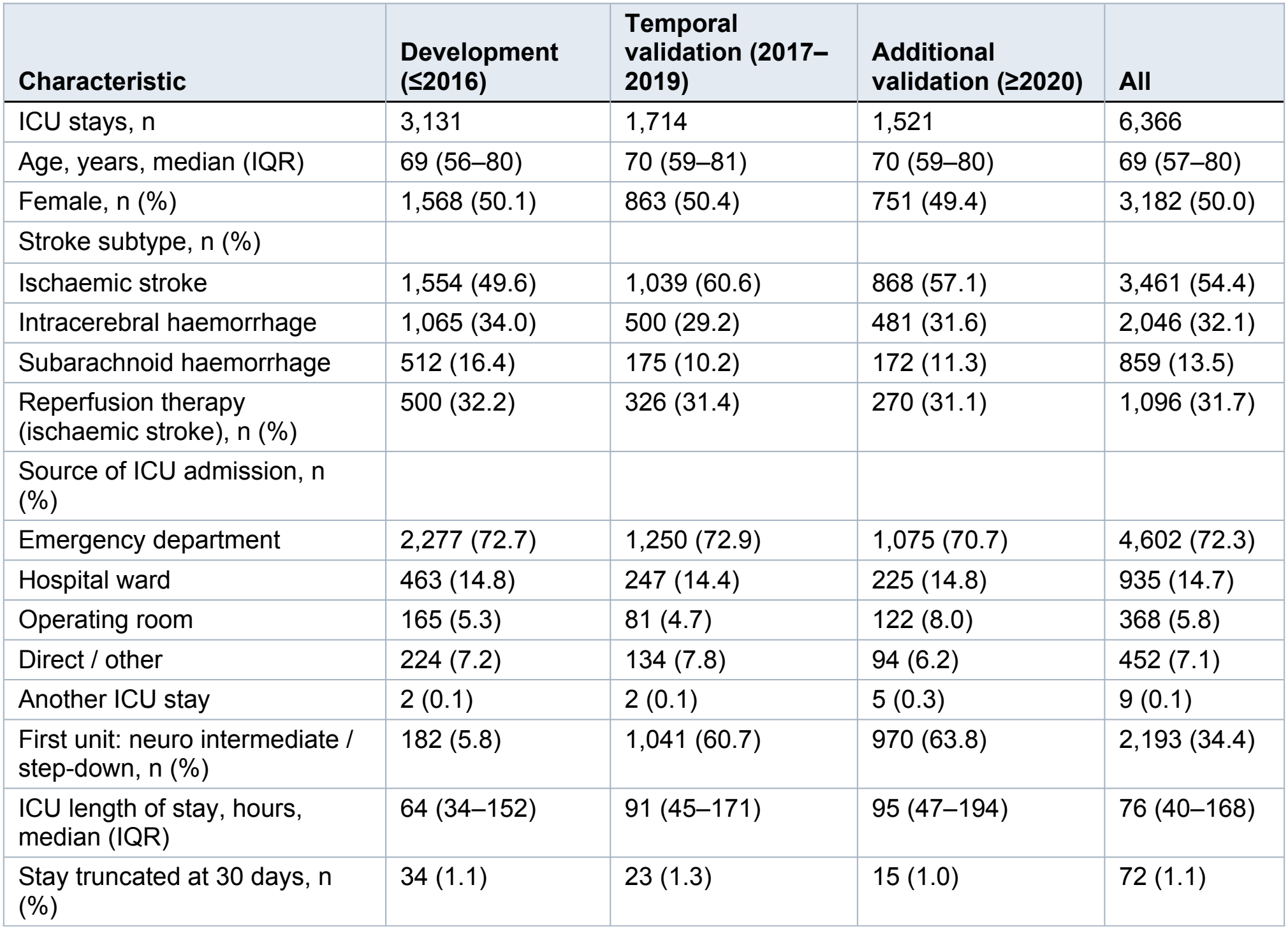
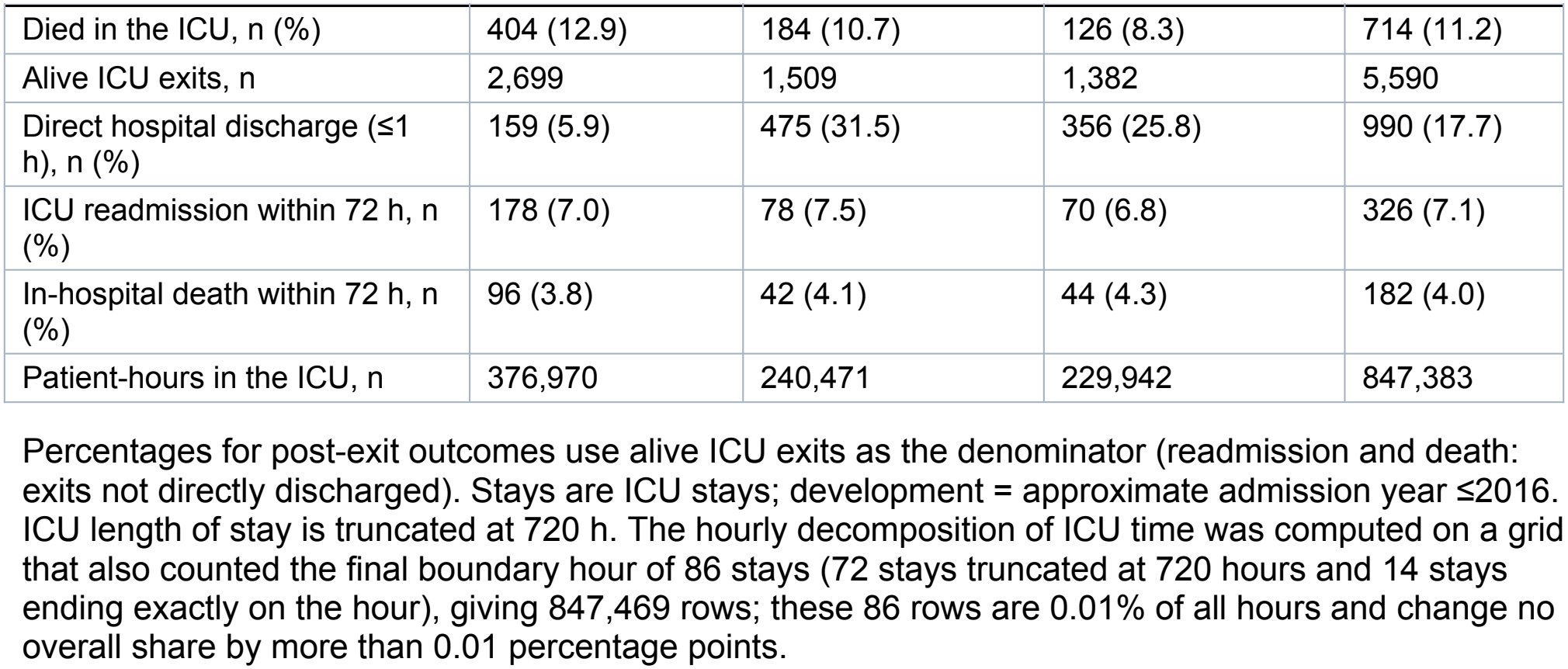
Characteristics of ICU stays by data split.

| Characteristic | Development (≤2016) | Temporal validation (2017–2019) | Additional validation (≥2020) | All |
|---|---|---|---|---|
| ICU stays, n | 3,131 | 1,714 | 1,521 | 6,366 |
| Age, years, median (IQR) | 69 (56–80) | 70 (59–81) | 70 (59–80) | 69 (57–80) |
| Female, n (%) | 1,568 (50.1) | 863 (50.4) | 751 (49.4) | 3,182 (50.0) |
| Stroke subtype, n (%) |  |  |  |  |
| Ischaemic stroke | 1,554 (49.6) | 1,039 (60.6) | 868 (57.1) | 3,461 (54.4) |
| Intracerebral haemorrhage | 1,065 (34.0) | 500 (29.2) | 481 (31.6) | 2,046 (32.1) |
| Subarachnoid haemorrhage | 512 (16.4) | 175 (10.2) | 172 (11.3) | 859 (13.5) |
| Reperfusion therapy (ischaemic stroke), n (%) | 500 (32.2) | 326 (31.4) | 270 (31.1) | 1,096 (31.7) |
| Source of ICU admission, n (%) |  |  |  |  |
| Emergency department | 2,277 (72.7) | 1,250 (72.9) | 1,075 (70.7) | 4,602 (72.3) |
| Hospital ward | 463 (14.8) | 247 (14.4) | 225 (14.8) | 935 (14.7) |
| Operating room | 165 (5.3) | 81 (4.7) | 122 (8.0) | 368 (5.8) |
| Direct / other | 224 (7.2) | 134 (7.8) | 94 (6.2) | 452 (7.1) |
| Another ICU stay | 2 (0.1) | 2 (0.1) | 5 (0.3) | 9 (0.1) |
| First unit: neuro intermediate / step-down, n (%) | 182 (5.8) | 1,041 (60.7) | 970 (63.8) | 2,193 (34.4) |
| ICU length of stay, hours, median (IQR) | 64 (34–152) | 91 (45–171) | 95 (47–194) | 76 (40–168) |
| Stay truncated at 30 days, n (%) | 34 (1.1) | 23 (1.3) | 15 (1.0) | 72 (1.1) |
| Died in the ICU, n (%) | 404 (12.9) | 184 (10.7) | 126 (8.3) | 714 (11.2) |
| Alive ICU exits, n | 2,699 | 1,509 | 1,382 | 5,590 |
| Direct hospital discharge (≤1 h), n (%) | 159 (5.9) | 475 (31.5) | 356 (25.8) | 990 (17.7) |
| ICU readmission within 72 h, n (%) | 178 (7.0) | 78 (7.5) | 70 (6.8) | 326 (7.1) |
| In-hospital death within 72 h, n (%) | 96 (3.8) | 42 (4.1) | 44 (4.3) | 182 (4.0) |
| Patient-hours in the ICU, n | 376,970 | 240,471 | 229,942 | 847,383 |

Percentages for post-exit outcomes use alive ICU exits as the denominator (readmission and death: exits not directly discharged). Stays are ICU stays; development = approximate admission year ≤2016. ICU length of stay is truncated at 720 h. The hourly decomposition of ICU time was computed on a grid that also counted the final boundary hour of 86 stays (72 stays truncated at 720 hours and 14 stays ending exactly on the hour), giving 847,469 rows; these 86 rows are 0.01% of all hours and change no overall share by more than 0.01 percentage points.

The analyses followed two statistical analysis plans, both finalised before the relevant results were produced and provided in the Supplement: one for the description of ICU time, amended three times with the reasons recorded, and one for the simulator. The first amendment, which changed the main presentation to separate results by unit type, was made after the descriptive results had been seen. Reporting follows TRIPOD+AI for prediction,[11] STRESS for simulation,[12] and RECORD for routinely collected data.[13]

### Operational definition of transfer readiness

We defined readiness to leave the ICU hour by hour from the literature, using only data available at each hour (Supplementary Methods S2).[1,14] Physiological readiness required four conditions together: no invasive or non-invasive ventilation or high-flow oxygen; no continuous vasopressor; no confirmed new decline in Glasgow Coma Scale (GCS), no continuous sedation or marked agitation, and no recorded seizure; and no critical metabolic abnormality or ICU-level corrective treatment.

Stroke-extended readiness added two stroke-specific conditions: no continuous intravenous antihypertensive infusion, and completion of the first 24 hours after reperfusion therapy.[15] Readiness was confirmed when the conditions had been met continuously for 6 hours, at the sixth hour. The definition captures conditions observable in the record; it is not the care team’s transfer decision.

### Hourly decomposition of ICU time

Each ICU hour was assigned to one of three categories: T1, physiological readiness not met (organ-support needs); T2, physiological readiness met but not the two stroke-specific conditions; and T3, both met (ICU time not explained by observed needs). T3 is not a measure of waiting: it may include monitoring, nursing needs and clinical judgement that the data do not capture. We report the share of hours (main measure) and the mean share per patient, by unit type, stroke subtype and day of stay, with 95% CIs from a patient-level bootstrap. Reproducibility across eras was judged against criteria set in advance.

Two analyses were added in amendment 1.2, before their results or any post-exit outcome data had been examined. First, monitoring intensity within T3: an hour was classed as high intensity if 3 or more GCS assessments were recorded in the preceding 6 hours. Second, criterion validity: among patients leaving the ICU alive, we compared three groups defined at exit (readiness met at the last hour; met earlier but lost; never met) for ICU readmission in the same hospitalisation (primary outcome), post-ICU in-hospital death, and their composite. We used logistic regression adjusted for age, sex, subtype, log ICU length of stay, unit type at exit, era, reperfusion therapy, and exit between 19:00 and 07:00.

### Simulator structure

The simulator advances each patient one hour at a time through a fixed sequence of modules (Figure 1, Table 2).

**Table 2.** Simulator modules, inputs and selected algorithms.

| Module | Predicts | Time step | Risk set | Selected algorithm |
|---|---|---|---|---|
| D2-0 | First condition state (4 HMM states) | Hour 6 | Alive in ICU at hour 6 | Logistic splines (L1) |
| D2 | Next condition state, one module per current state: renal dysfunction / respiratory support / impairment with low support / preserved | Every 6 h | Alive in ICU | L2 / L3 / L1 / L2 |
| D1 | Death in the ICU | Hourly | Alive in ICU | L1 |
| C1 | Stay / move to other unit type / leave ICU | Hourly | Alive in ICU | L1 (no candidate qualified) |
| D3a | Readiness gained (unmet → met) | Hourly | Readiness not met | LightGBM (L3) |
| D3b | Readiness lost (met → unmet) | Hourly | Readiness met | L3 |
| P1a | Direct hospital discharge (≤1 h) | At exit | Alive exits | L1 (no candidate qualified) |
| P1b | ICU readmission / in-hospital death / discharge | Hourly to 72 h | Alive exits not directly discharged | L3 |

Inputs for all ICU modules: condition state; readiness met now, ever confirmed, hours since confirmation; hours in ICU; unit type; hour of day (4-hour blocks); weekend; age; stroke subtype; reperfusion therapy. Module-specific formulas are in Supplementary Table S5. L2 modules selected the strongest penalty in the grid.

**Figure 1.**
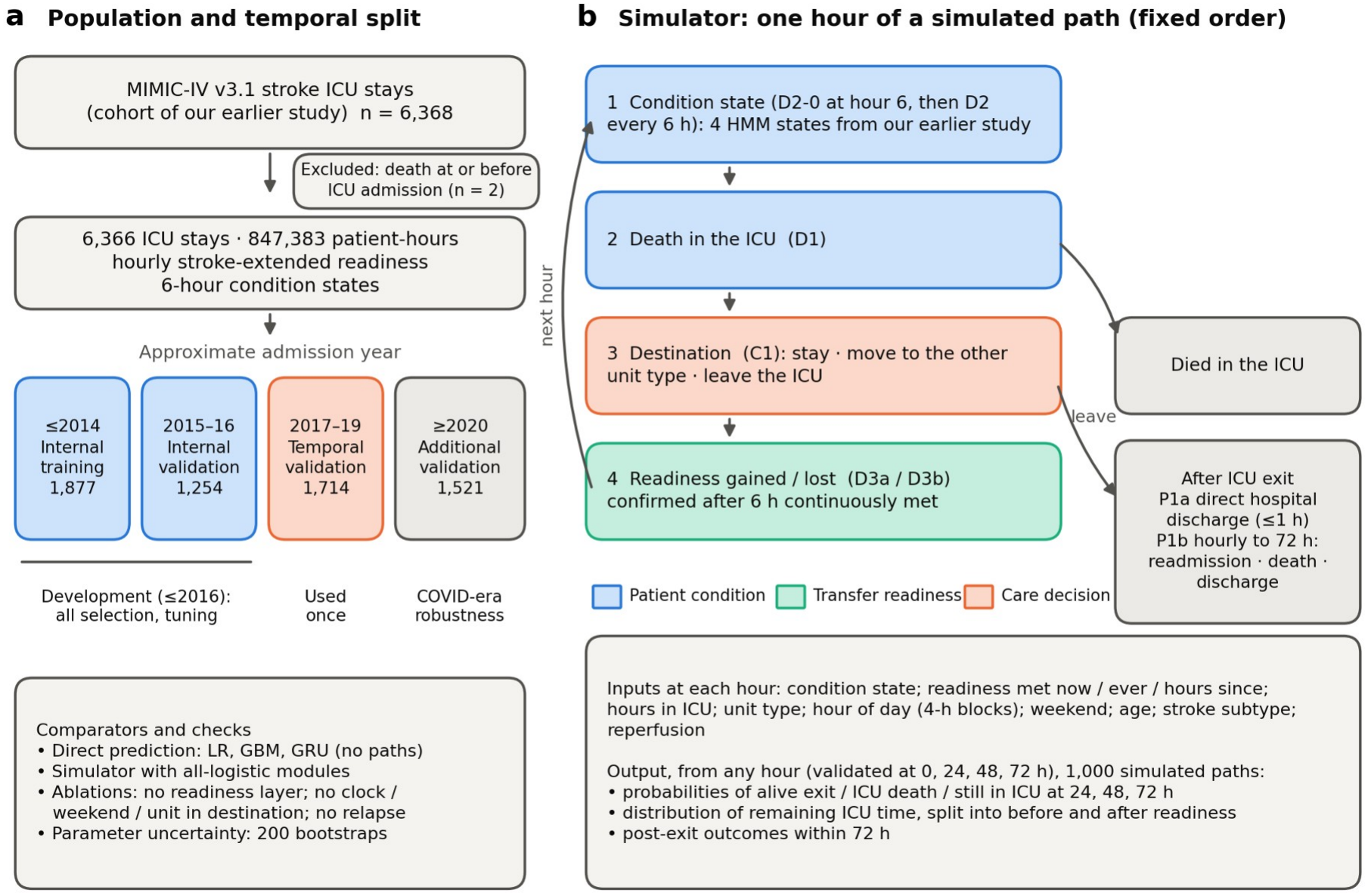
Study design and simulator structure. (a) Study population and temporal split: the stroke ICU cohort of our earlier study, the exclusions, and the four sets defined by approximate admission year (internal training up to 2014, internal validation 2015–2016, temporal validation 2017–2019 and additional validation from 2020), with the comparators and checks. (b) Simulator: the fixed order of modules in each simulated hour (condition state, death in the ICU, destination and readiness), coloured by layer (patient condition, transfer readiness, care decision) and leading to death in the ICU or to post-exit outcomes, with the simulator’s inputs and outputs below. Alt text: Two-panel diagram. The left panel shows 6,366 ICU stays divided into development (up to 2016), temporal validation (2017–2019) and additional validation (2020 onwards). The right panel shows the hourly loop of condition state, death, destination and readiness modules, leading to death in the ICU or to post-exit outcomes.

- **Condition state (D2-0, D2):** one of four states from a hidden Markov model frozen in our earlier study.[10] The states are neurologically preserved with low support; neurological impairment with low support; neurological impairment with renal dysfunction; and neurological impairment with respiratory support. The first state is assigned at hour 6 and updated every 6 hours, using forward-filtered (past-only) probabilities.
- **Death in the ICU (D1):** hourly.
- **Destination (C1):** each hour, stay, move to the other unit type, or leave the ICU, modelled as competing outcomes.
- **Readiness gained or lost (D3a, D3b):** hourly.
- **After exit:** direct hospital discharge within 1 hour (P1a), and hourly outcomes for 72 hours (P1b: ICU readmission, in-hospital death, discharge).

Inputs are listed in Table 2. From any starting hour, the simulator generated 1,000 paths per patient. From these it estimated probabilities of alive exit, ICU death and remaining in the ICU at 24, 48 and 72 hours, the distribution of remaining ICU time split into time before and after readiness, and post-exit outcomes.

### Temporal split

MIMIC-IV shifts dates for privacy. We approximated the admission year from the anchor year group and the offset between admission and anchor years (precision about ±1 year). Development used stays admitted up to 2016 (internal training up to 2014, n=1,877; internal validation 2015–2016, n=1,254). Temporal validation used 2017–2019 (n=1,714) and was run once. Stays from 2020 onwards (n=1,521) formed an additional validation set covering the COVID-19 period.

### Algorithms, tuning and selection

Each module had three candidate algorithms, with all hyperparameter grids fixed in advance and not extended after validation:

- **L1:** logistic regression with natural cubic splines.
- **L2:** elastic net with pre-listed interactions.
- **L3:** LightGBM,[16] with separate cross-fitting folds for early stopping and calibration.

Folds were grouped by patient. Selection on the internal validation period was deterministic. Among candidates with a calibration slope of 0.8–1.2, we chose the one with the lowest log loss. If a more interpretable candidate was within 1% of it, we chose that one instead. If no candidate qualified, we used L1. At the system level, configuration A (per-module choices) was compared with configuration B (all L1) on five required checks of simulated paths. A would be retained unless its Brier score was more than 2% worse. Final modules were refitted on all development data with fixed hyperparameters and frozen.

### Comparators and evaluation

Three direct models predicted the same outcomes from each landmark without generating paths:

- logistic regression and gradient boosting (GBM), both in discrete-time competing-risk form;
- a gated recurrent unit network using the preceding 72 hours.

An empirical baseline gave each patient the development-set rates for their stroke subtype and landmark (Supplementary S7). The strongest model was chosen on internal validation by log loss (GBM).

The single primary comparison was the Brier score of the 24-hour probability of leaving the ICU alive, pooled over landmarks at 0, 24, 48 and 72 hours. We compared the simulator (configuration A) with the strongest direct model, with a 95% CI from 1,000 patient-level bootstrap samples. The key secondary measure was the Wasserstein distance between simulated and observed distributions of ICU length of stay. Other secondary measures were:

- Brier score, calibration slope and C statistic for alive exit and ICU death at 24, 48 and 72 hours;
- remaining ICU time and the coverage of its 80% prediction interval;
- results by unit type and subtype;
- post-exit predictions;
- logical consistency checks of simulated paths.

Parameter uncertainty was estimated from 200 bootstrap refits of the development data; it differs from the 80% prediction intervals, which describe uncertainty in an individual’s future path.

We tested each layer by refitting the simulator without it, and re-selected the changed modules by the same rule:

- **(A)** no readiness inputs in the destination and post-exit modules;
- **(B)** no hour of day, weekend or unit type in the destination module;
- **(C)** no loss of readiness after confirmation.

Pre-specified sensitivity analyses repeated the full pipeline (1) with the split based on anchor year groups, and (2) with physiological in place of stroke-extended readiness. A third repeated the uncertainty analysis with 100 bootstrap samples.

### Interpretability

For L1 and L2 modules we report odds ratios and effect plots. For L3 modules we computed TreeSHAP values[17] and checked whether their direction agreed with the matching L1 term (Spearman correlation). At the individual level we simulated paths for three fictional patient profiles, and split remaining time into time to readiness and time from readiness to exit. Each prediction carries applicability flags (Supplementary Table S14).

### Software

Analyses used Python 3.9 with pandas, scikit-learn 1.6.1, LightGBM 4.6.0 and PyTorch 2.8.0. The use of artificial intelligence tools is described in the Acknowledgements.

## RESULTS

### Population

Table 1 shows the 6,366 stays by split. Median age was 69 years; 50.0% were women; 54.4% had ischaemic stroke, 32.1% intracerebral haemorrhage and 13.5% subarachnoid haemorrhage. Care structure changed between eras. The first unit was a neuro intermediate unit in 5.8% of development stays but in 60.7% of 2017–2019 stays. Direct hospital discharge from an ICU-level unit rose from 5.9% to 31.5% of alive exits. Median ICU length of stay rose from 64 to 91 hours.

### Where ICU time goes

Over all stays, 46.9% of ICU hours were T1, 4.2% T2 and 49.0% (95% CI 47.7–50.2) T3. The split differed sharply by unit type (Figure 2): T1 and T3 made up 61.4% and 34.1% (33.0–35.3) of hours in standard ICUs, and 9.0% and 87.6% (86.7–88.5) in neuro intermediate units. In standard ICUs, the T1 share rose with length of stay, from 53% on day 1 to 73% on days 15–30. T2 was concentrated on day 1 (16% of hours), reflecting monitoring after reperfusion and antihypertensive infusions.

**Figure 2.**
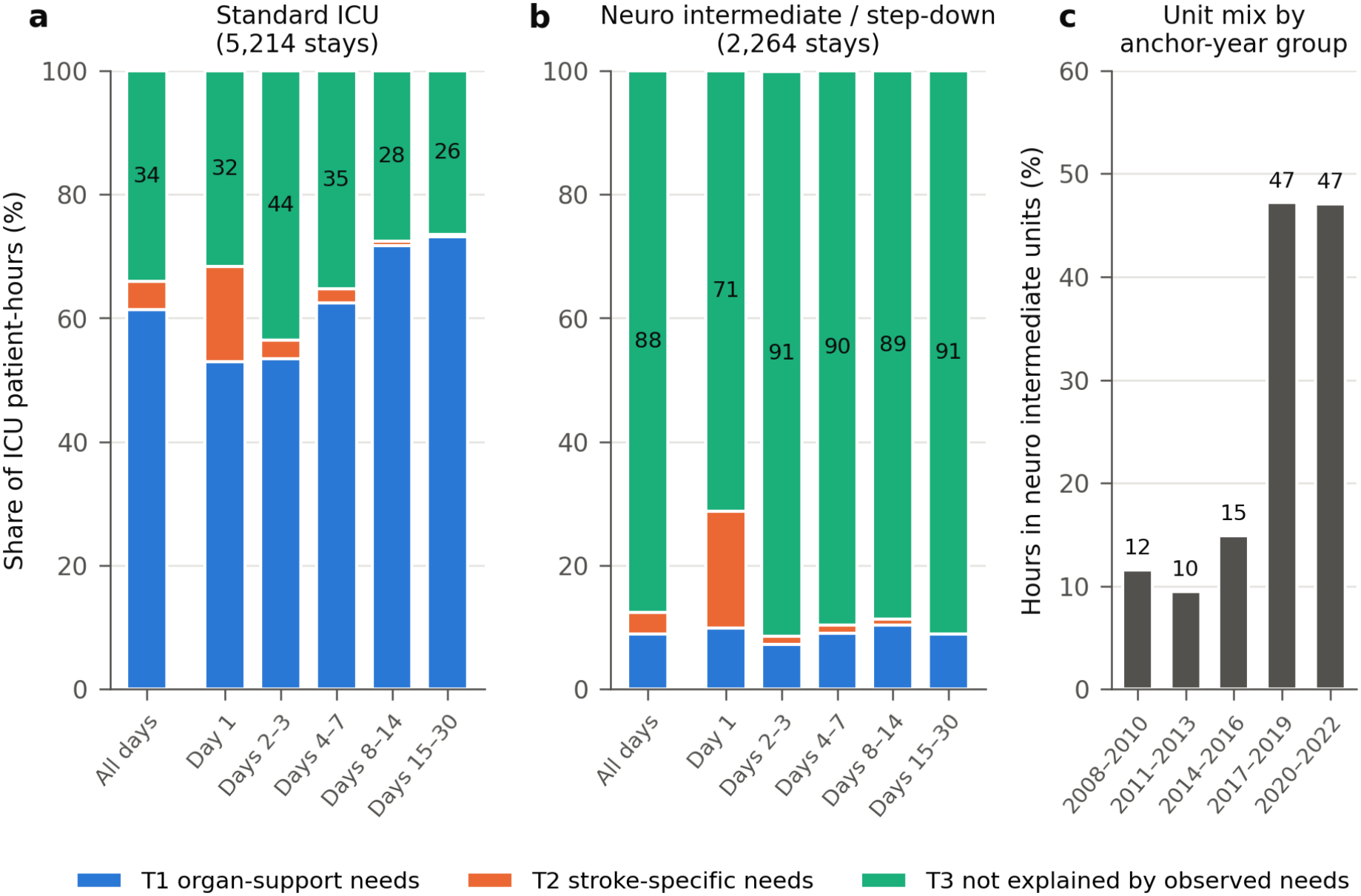
Decomposition of ICU time. Share of patient-hours in T1 (organ-support needs), T2 (stroke-specific needs) and T3 (not explained by observed needs), in (a) standard ICUs and (b) neuro intermediate or step-down units, overall and by day of stay; (c) share of ICU hours spent in neuro intermediate units by anchor year group. Alt text: Stacked bar charts. In standard ICUs, T3 is about one third of hours; in intermediate units, about 90%. A third panel shows the share of hours in intermediate units rising from 10–15% before 2017 to 47% afterwards.

By the pre-specified criteria, the overall T3 share was not stable across eras. It was 40.2–41.2% in 2008–2016, 57.1% in 2017–2019, and 62.3% in 2020–2022. In a post hoc analysis this change followed the unit mix: the share of hours spent in intermediate units rose from 9.5–14.9% to 47.1– 47.2%. Within each unit type, T3 was stable across eras (standard ICU 30.0–38.1%; intermediate 84.1– 89.6%).

Much of T3 involved frequent neurological assessment. In 54.2% of T3 hours, 3 or more GCS assessments had been recorded in the preceding 6 hours (standard ICU 59.1%; intermediate 49.2%). This share fell with length of stay, from 66–70% on day 1 to 22–24% on days 15–30.

### Readiness dynamics

The cumulative incidence of reaching stroke-extended readiness was 46.4% by 24 hours and 72.8% by 72 hours. It was faster for stays that started in an intermediate unit (64.4% by 24 hours) than in a standard ICU (37.3%) (Figure 3).

**Figure 3.**
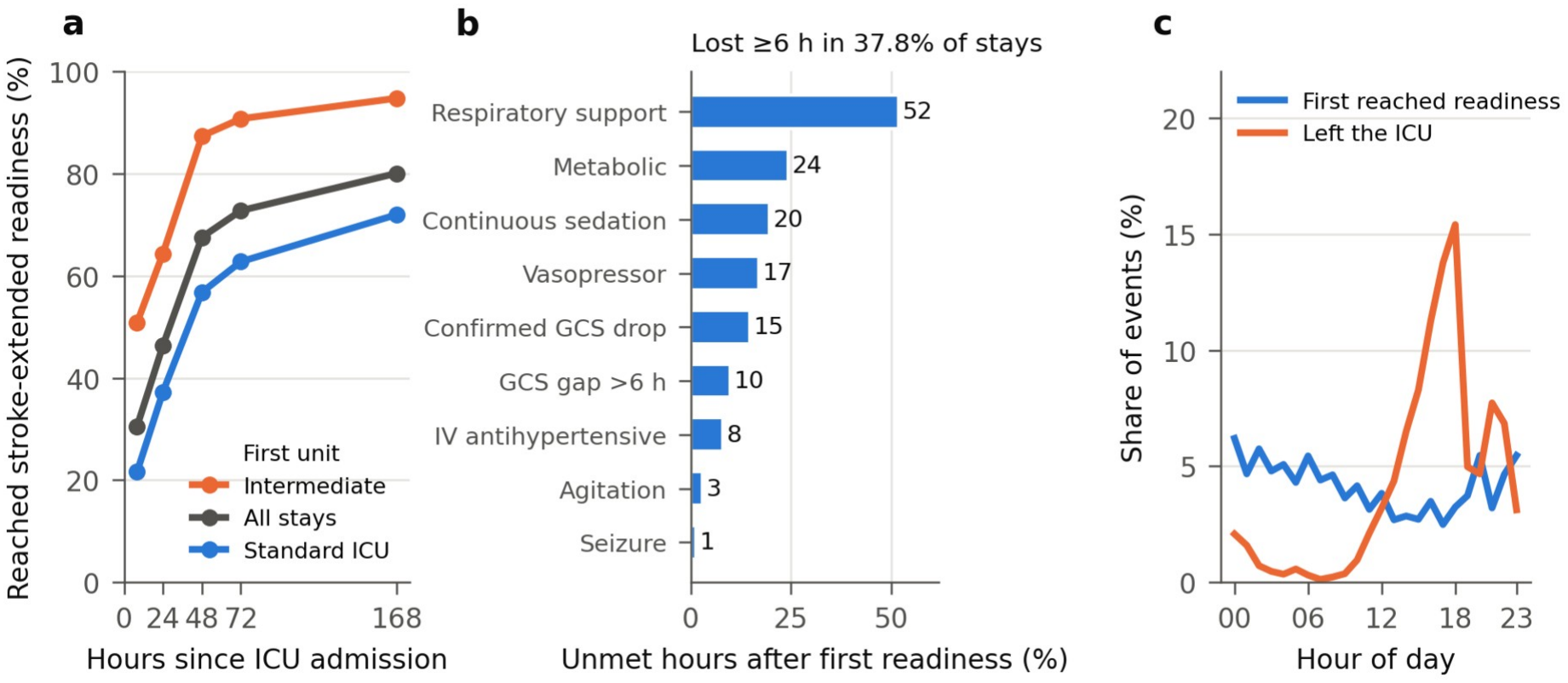
Transfer readiness over time. (a) Cumulative incidence of reaching stroke-extended readiness, with death as a competing event, by first unit type. (b) Criteria unmet in hours after first readiness, among stays that lost readiness for 6 hours or more (37.8%). (c) Hour of day at which readiness was first reached and at which patients left the ICU. Alt text: Line chart showing faster readiness in intermediate units; bar chart showing respiratory support as the most common reason readiness was lost; line chart showing readiness spread across the day while exits peak at 18:00.

Among 4,905 patients who left alive after reaching readiness, the median time from readiness to exit was 53.7 hours (IQR 21.4–121.4). Only 4.6% left within 6 hours.

Readiness was often lost again: 37.8% of stays had at least one run of 6 or more consecutive hours with criteria unmet after first confirmed readiness. The criteria most often unmet in these hours were respiratory support (52%), metabolic abnormality (24%) and continuous sedation (20%). This reflects a change in readiness status, not necessarily deterioration.

Readiness was first reached throughout the day, whereas exits clustered between 14:00 and 19:00. Yet 34.2% of exits occurred between 19:00 and 07:00.

Readiness status at exit was associated with post-ICU death but not with ICU readmission (Table 3). Post-ICU in-hospital death was 3.3% in patients who met readiness at exit, 13.6% in those who had lost it, and 27.0% in those who never met it (adjusted odds ratios 3.40 [2.48–4.66] and 8.54 [6.55–11.13]). Readmission was 9.8%, 11.0% and 11.4% (0.96 [0.71–1.29] and 1.07 [0.81–1.40]). Excluding 36 patients with a comfort-measures-only order before exit did not change these results.

**Table 3.**
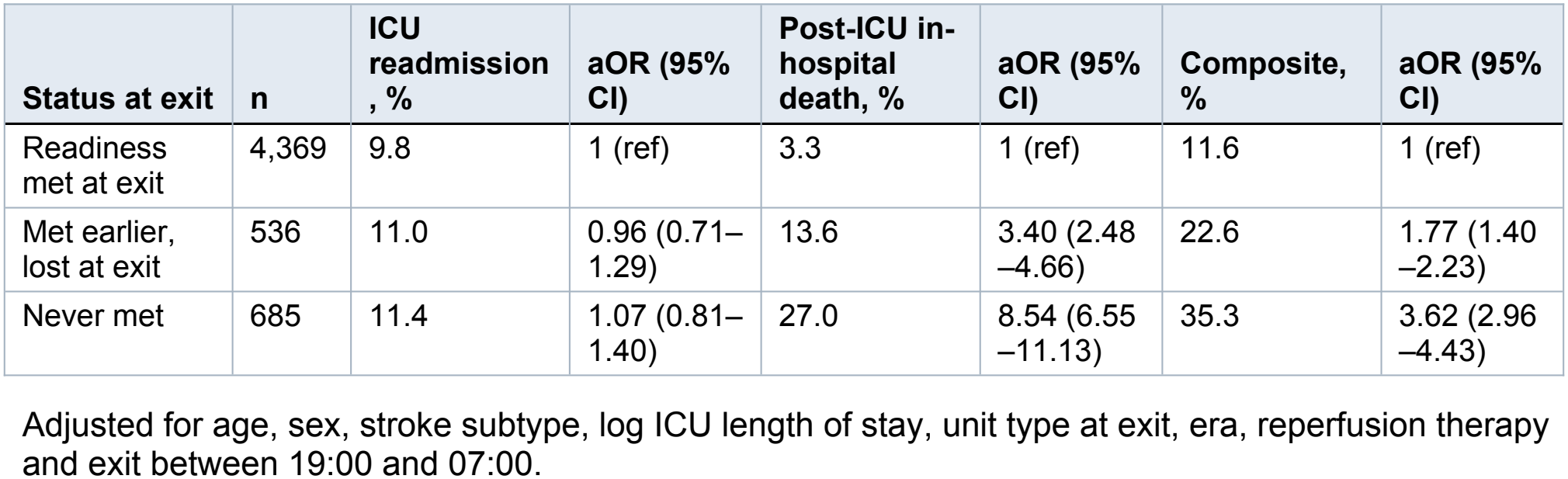
Readiness status at ICU exit and post-exit outcomes (alive exits, n=5,590).

| **Status at exit** | **n** | **ICU readmission, %** | **aOR (95% CI)** | **Post-ICU in-hospital death, %** | **aOR (95% CI)** | **Composite, %** | **aOR (95% CI)** |
|---|---|---|---|---|---|---|---|
| Readiness met at exit | 4,369 | 9.8 | 1 (ref) | 3.3 | 1 (ref) | 11.6 | 1 (ref) |
| Met earlier, lost at exit | 536 | 11.0 | 0.96 (0.71–1.29) | 13.6 | 3.40 (2.48–4.66) | 22.6 | 1.77 (1.40–2.23) |
| Never met | 685 | 11.4 | 1.07 (0.81–1.40) | 27.0 | 8.54 (6.55–11.13) | 35.3 | 3.62 (2.96–4.43) |

Adjusted for age, sex, stroke subtype, log ICU length of stay, unit type at exit, era, reperfusion therapy and exit between 19:00 and 07:00.

### Simulator development

Table 2 shows the selected algorithms: LightGBM for both readiness transitions, one state transition and post-exit outcomes; elastic net, at the strongest penalty, for two state transitions; logistic splines elsewhere. No candidate met the calibration criterion for the destination module (C1) or for direct discharge (P1a), so L1 was used for both, as the rule required.

This failure was anticipated in the implementation log before validation. Moves between unit types occurred only 3 times in the internal training period, compared with 121 times in 2015–2016 and 618 times in 2017–2019. Both configurations passed all system checks (internal Brier A 0.1261, B 0.1260), so A was retained; no simulated path had a logical violation.

### Temporal validation

In 2017–2019 (5,543 patient-landmarks), the simulator and GBM were indistinguishable on the primary measure (Table 4):

**Table 4.** Temporal validation (2017–2019) and additional validation (2020 onwards).

|  | Simulator A | Simulator B (all L1) | LR | GBM | GRU |
|---|---|---|---|---|---|
| ***2017–2019: 1,714 stays, 5,543 patient-landmarks*** |  |  |  |  |  |
| **Primary: 24-h alive-exit Brier** | **0.1238** | 0.1245 | 0.1237 | **0.1245** | 0.1347 |
| 24-h alive-exit C statistic | 0.708 | 0.708 | 0.718 | 0.740 | 0.749 |
| 24-h alive-exit calibration slope | 0.95 | 1.00 | 1.04 | 0.84 | 0.84 |
| 24-h alive exit, predicted − observed | 0.077 | 0.083 | 0.082 | 0.081 | 0.122 |
| 48-h alive-exit Brier | 0.203 | 0.207 | 0.210 | 0.207 | 0.214 |
| 48-h calibration slope | 0.89 | 0.91 | 0.75 | 0.76 | 0.84 |
| 72-h alive-exit Brier | 0.223 | 0.230 | 0.230 | 0.228 | 0.231 |
| 72-h calibration slope | 1.01 | 1.00 | 0.69 | 0.75 | 0.86 |
| 24-h ICU-death C statistic | 0.837 | 0.828 | 0.809 | 0.861 | 0.852 |
| 72-h ICU-death Brier | 0.0429 | 0.0429 | 0.0429 | 0.0417 | 0.0421 |
| LOS Wasserstein, full stay (h) | 30.4 | 38.4 | — | — | — |
| LOS Wasserstein, to 72 h (h) | 6.60 | 6.96 | 7.85 | 6.14 | — |
| ***2020 onwards: 1,521 stays, 4,979 patient-landmarks*** |  |  |  |  |  |
| **Primary: 24-h alive-exit Brier** | **0.1261** | 0.1267 | 0.1245 | **0.1260** | 0.1390 |
| 24-h alive-exit C statistic | 0.684 | 0.687 | 0.711 | 0.741 | 0.742 |
| 24-h alive-exit calibration slope | 0.85 | 0.91 | 1.02 | 0.85 | 0.83 |
| 24-h alive exit, predicted − observed | 0.092 | 0.098 | 0.096 | 0.100 | 0.139 |
| 48-h alive-exit Brier | 0.208 | 0.212 | 0.218 | 0.215 | 0.224 |
| 48-h calibration slope | 0.81 | 0.85 | 0.71 | 0.72 | 0.79 |
| 72-h alive-exit Brier | 0.232 | 0.240 | 0.247 | 0.242 | 0.246 |
| 72-h calibration slope | 0.97 | 0.98 | 0.64 | 0.69 | 0.81 |
| 24-h ICU-death C statistic | 0.805 | 0.808 | 0.799 | 0.828 | 0.826 |
| 72-h ICU-death Brier | 0.0329 | 0.0328 | 0.0330 | 0.0335 | 0.0324 |
| LOS Wasserstein, full stay (h) | 41.6 | 49.6 | — | — | — |
| LOS Wasserstein, to 72 h (h) | 7.92 | 8.26 | 9.45 | 7.40 | — |

Primary comparison, simulator A − GBM (95% CI, 1,000 patient-level bootstrap samples): 2017–2019, −0.0007 (−0.0030 to 0.0016); 2020 onwards, +0.0002 (−0.0026 to 0.0027). GBM was the strongest direct model on internal validation. Direct models predict only to 72 hours, so the full-stay distance is given for the simulators only. All models were fitted on stays up to 2016 and frozen before validation.

- Brier score: 0.1238 vs 0.1245.
- Difference: −0.0007 (95% CI −0.0030 to 0.0016).

This shows neither superiority nor equivalence. The empirical baseline scored 0.1249 despite a C statistic of 0.63: the 24-hour Brier score mainly reflected the overall level of exit, which every model overpredicted.

At 48 and 72 hours, the simulator had the lowest Brier scores (0.203 and 0.223) and calibration slopes closest to 1 (0.89 and 1.01, against 0.69–0.86 for direct models; Figure 4). Direct models discriminated better: the C statistic for 24-hour alive exit was 0.74–0.75 for GBM and GRU and 0.71 for the simulator. Better calibration did not mean better ranking.

**Figure 4.**
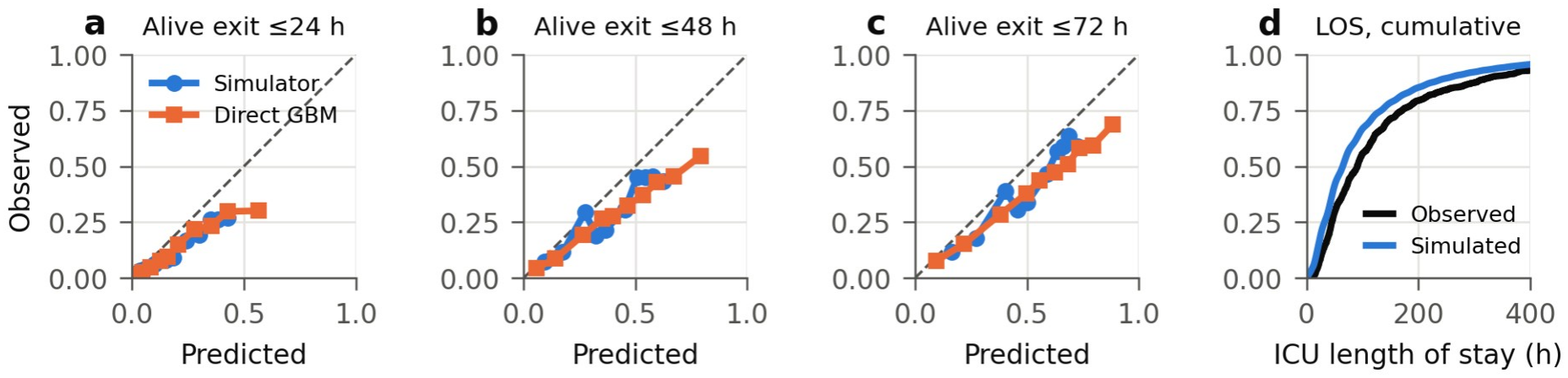
Temporal validation, 2017–2019. (a–c) Calibration of the predicted probability of leaving the ICU alive within 24, 48 and 72 hours for the simulator and the gradient boosting model, by deciles of predicted risk. (d) Cumulative distribution of observed and simulated ICU length of stay. Alt text: Three calibration plots in which the simulator lies closer to the diagonal than gradient boosting at 48 and 72 hours, and both lie below it; and a cumulative curve in which simulated stays are shorter than observed.

For ICU death, Brier scores were similar; 24-hour C statistics were 0.84 (simulator) and 0.86 (GBM).

Over the full stay, per-module choices reproduced the length-of-stay distribution better than all-logistic modules (Wasserstein distance 30.4 vs 38.4 hours). Up to 72 hours, where direct models can be compared, GBM was closer (6.14 vs 6.60). The 80% prediction intervals for remaining ICU time covered 81% of alive exits and 75% of ICU deaths.

All models overpredicted ICU exit, by 0.08–0.12 in absolute probability at 24 hours, and the simulated median stay was 65 hours against 91 observed. The overprediction was larger than the parameter uncertainty (half-width of the 95% interval 0.03–0.045 for alive exit), so it was systematic rather than due to limited data.

The simulator located the error. Among alive exits in 2017–2019, the median time from admission to readiness was 12 hours observed and 15 hours simulated. The median time from readiness to exit was 68 hours observed and only 40 hours simulated. The time before readiness was reproduced well; the error lay after readiness. The destination module was estimated mainly from years with a different unit mix, which may be one source of the error. This is a model diagnosis; it does not show that any real clinical decision caused the error.

The direct-discharge module (P1a) failed in the same way: it predicted 5.9% against 31.5% observed (C statistic 0.54). Post-ICU in-hospital death was well discriminated (C statistic 0.94), and ICU readmission moderately (0.63).

In the 2020 onwards validation set (4,979 patient-landmarks), conclusions were the same. The primary difference was +0.0002 (−0.0026 to 0.0027). The simulator again had the lowest 48- and 72-hour Brier scores and the best 72-hour calibration (slope 0.97 vs 0.64–0.81), while GBM again discriminated better (0.74 vs 0.68). The empirical baseline had the lowest 24-hour Brier score of all (0.1213 vs 0.1245 –0.1390).

### Contribution of each layer

Removing any layer worsened the primary measure, with 95% CIs excluding zero (Figure 5; Supplementary Table S8). Without readiness inputs the Brier score rose by 0.0018 (0.0002–0.0034), mainly through lost discrimination (C statistic 0.708 to 0.668). Without hour of day, weekend and unit in the destination module it rose by 0.0047 (0.0038–0.0055), through worse overprediction of exit. Without readiness relapse it rose by 0.0023 (0.0016–0.0029), and the length-of-stay distance grew from 30.4 to 35.9 hours.

**Figure 5.**
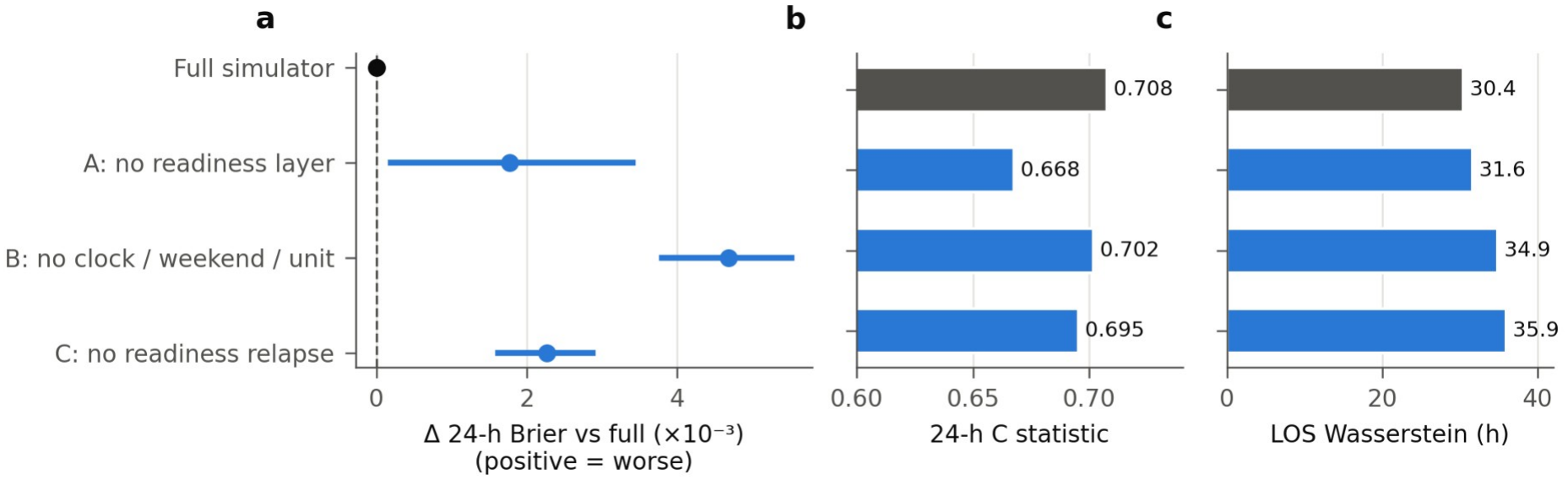
Contribution of each layer. (a) Difference in 24-hour alive-exit Brier score between each reduced simulator and the full simulator, with 95% CIs. (b) C statistic. (c) Wasserstein distance between simulated and observed length of stay. Alt text: Forest plot in which all three reduced simulators are worse than the full simulator; bar charts showing the largest loss of discrimination without the readiness layer and the largest loss in length-of-stay fit without relapse.

### Interpretability

The destination module expressed a clear rule (Figure 6a). For ready patients in a standard ICU, the probability of leaving in the next hour peaked at 7.7% between 16:00 and 20:00 and was near zero between 04:00 and 08:00. For patients not ready, it was about ten times lower at every hour. This pattern was learned from past records and is not advice on when to transfer.

**Figure 6.**
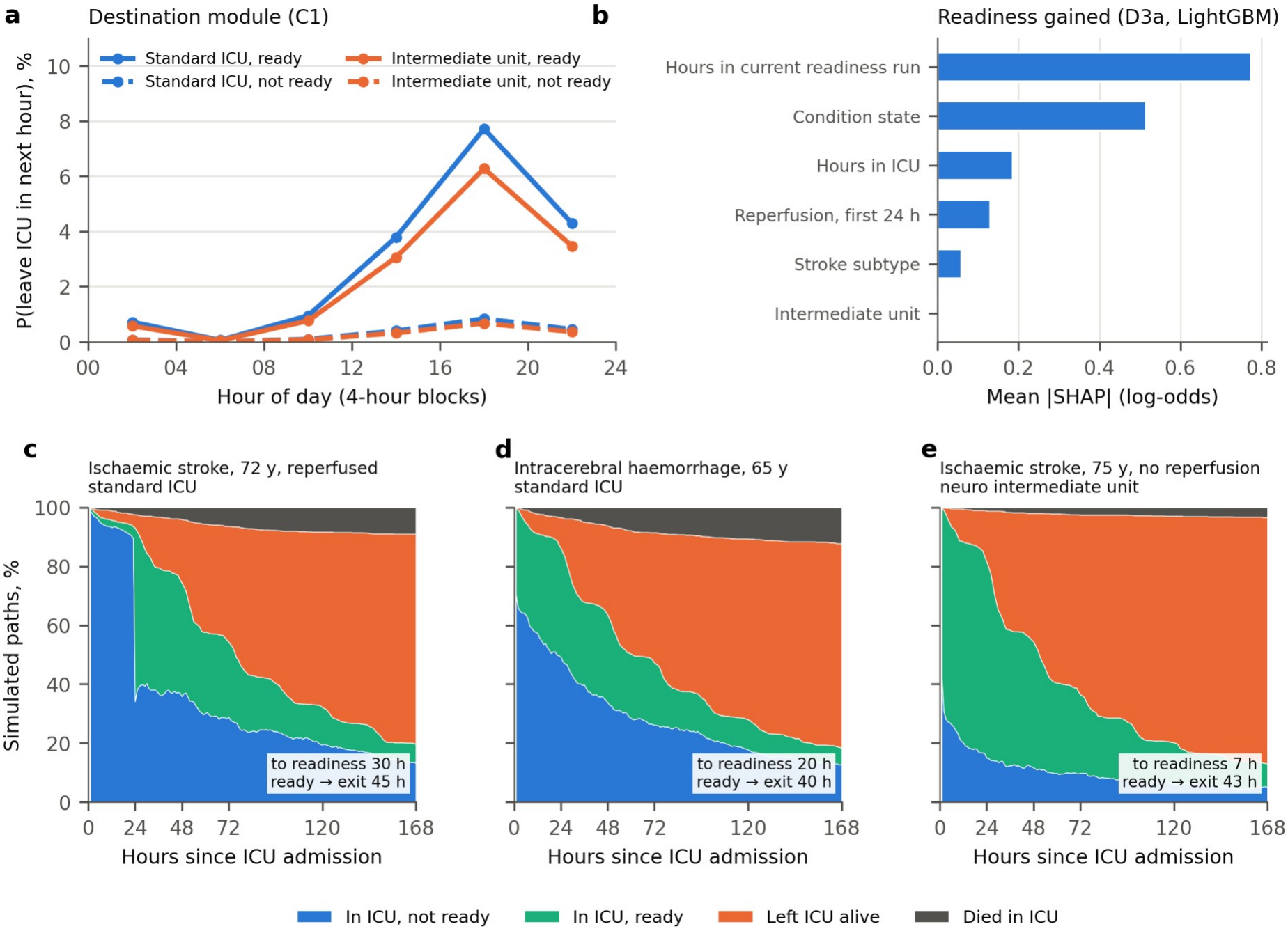
Interpretability. (a) Destination module: probability of leaving the ICU in the next hour by hour of day, unit type and readiness. (b) Mean absolute SHAP values in the readiness-gained module (LightGBM). (c–e) Simulated paths for three fictional patient profiles (not real patients), showing the share of 1,000 paths in the ICU before readiness, in the ICU after readiness, after alive exit and after ICU death; the boxes give median times to readiness and from readiness to exit. Alt text: A line chart showing that the probability of leaving for ready patients is highest between 16:00 and 20:00; a bar chart showing that hours in the current readiness run and condition state contribute most to predicted readiness; and three stacked area charts of simulated paths over 7 days.

In the LightGBM modules, the main SHAP contributors were plausible, and each variable acted in the same direction as in the logistic version (Spearman 0.18–0.98; 0.55–0.98 for the leading variable of each module) (Figure 6b, Supplementary Figure S2). The one exception was log ICU length of stay in the post-exit module (−0.08).

In the three fictional profiles (Figure 6c–e), for a 72-year-old with reperfused ischaemic stroke in a standard ICU, the simulated median time to readiness was 30 hours and from readiness to exit 45 hours. These are model-generated paths, not predictions for real patients.

### Sensitivity analyses

With the split by anchor year group, the strongest direct model was GRU, and the simulator was again indistinguishable from it (difference −0.0000; −0.0015 to 0.0015). Its calibration advantage held, but GBM had the lowest 48- and 72-hour Brier scores (0.191 and 0.216 vs 0.195 and 0.222), so the 48–72-hour accuracy advantage was not reproduced under this split. With physiological readiness, GRU was again strongest (difference −0.0013; −0.0034 to 0.0009) and all secondary conclusions held. With 100 bootstrap samples, interval widths changed by about 5%.

## DISCUSSION

First, about half of ICU hours after stroke were not explained by observed clinical needs. This share differed sharply by unit type: one third of hours in standard ICUs, nearly nine in ten in neuro intermediate units.

Second, readiness was unstable: more than a third of stays lost it for 6 or more consecutive hours after first confirmation, and patients who left without meeting it had higher post-ICU mortality.

Third, modelling these layers separately generated complete paths. The simulator showed no clear difference from the strongest direct model on the primary measure, was better calibrated at longer horizons, and, when later data were systematically miscalibrated, gave clues to the source.

### ICU time and readiness

T3 should not be read as waiting time. More than half of T3 hours had recorded neurological assessments at least every 2 hours. Such monitoring may be the reason a patient stays in an ICU-level unit, particularly an intermediate unit, which exists to provide it. T3 is better read as ICU time that routine data do not explain: it marks the limit of the data, not the extent of wasted resources.

One-time criteria miss how often readiness is lost. Readiness status at exit was associated with post-ICU death but not with readmission. The definition thus captures part of clinical risk, but not the reasons for readmission. Patients who never met readiness include those leaving for palliative care, so the association should not be read as harm from early transfer, used to judge whether a transfer was premature, or used to infer the effect of earlier or later transfer.

### Simulator and direct prediction

Direct models ranked patients better. The simulator generated full paths and kept its calibration advantage across both validation sets and both sensitivity analyses; its 48–72-hour Brier advantage did not.

The two approaches answer different questions: a direct model is better for ranking which patients will leave soon, a simulator for questions about paths, such as how much time is spent before and after readiness, or what an hour-of-day pattern implies for occupancy. Each layer adds a small, stable gain: readiness mainly separates patients; timing mainly sets the overall level. These remain associations in past data, not predictions of what would happen if bed management changed.

### Detecting changes in care structure

The layered design was most useful when it failed: every model overpredicted exit after 2016. By comparing time to readiness with time from readiness to exit, the simulator suggested that the error lay mainly in the second interval, together with a marked underprediction of direct discharge, rather than in the patient’s condition or readiness. This matched the documented expansion of neuro intermediate care (Table 1). Whether the hospital changed its units or its recording practice cannot be determined from the data, and the error cannot be attributed to any specific clinical decision. When care changes, performance should be checked at each step of the path, which a single-output model cannot do. The planned external validation in the eICU Collaborative Research Database will test whether the condition and readiness layers transfer across hospitals.

### Comparison with previous work

ICU length-of-stay models have reported modest accuracy and poor transfer between settings.[3,4] Readiness has usually been assessed once, or by a team’s judgement recorded in whole days;[2,18] here it was measured hourly, and 37.8% of stays lost it for 6 or more consecutive hours after first confirmation. Discrete-event simulation in healthcare has mostly addressed operational efficiency;[5] the present simulator uses patient-level states and was validated on individual predictions. Its value lies in a testable representation of paths, not in reproducing clinical decisions in full.

### Limitations

This study has several limitations. It comes from a single centre whose unit structure changed over time, and the frozen model was miscalibrated in later years, so it should not be deployed before multicentre validation and updating. Admission years were approximated to within about one year. The readiness definition uses only recorded variables: ordered monitoring frequency, nursing judgement, bed availability and family wishes are not captured, and GCS and code-status records are sparse, so meeting the operational criteria is not equivalent to the care team’s judgement. The condition states were taken from a previously published model rather than re-derived, and the simulator represents neither treatment trajectories nor second ICU stays. The direct-discharge module failed in validation.

Differences between models were small, an empirical baseline scored as well or better on the primary measure, and the 48–72-hour accuracy advantage did not hold under the alternative split, so the simulator cannot be described as superior overall. The presentation by unit type was adopted after the descriptive results had been seen. Finally, the decompositions, layer removals and outcome associations describe patterns in past data, not causal effects, and do not support changes to transfer timing or policy.

## CONCLUSION

In ICU stays for stroke, about half of ICU time was not explained by observed clinical needs. This share depended largely on neuro intermediate care, and because records do not capture all monitoring needs and clinical judgement, it is not equivalent to unnecessary stay. More than a third of stays lost readiness for 6 or more consecutive hours after first confirmation. A simulator that keeps patient condition, readiness and care decisions separate showed no clear difference from the strongest direct model for 24-hour ICU exit, generated complete paths, and showed at which stage predictions drifted in later years. The results may inform evaluation of ICU transfer processes, but the model needs external validation and should not yet be a primary basis for transfer decisions. The frozen simulator and its code are available for external validation.

## Supporting information

Supplementary material S3: statistical analysis plans

## Data Availability

MIMIC-IV and MIMIC-IV-ED are available through PhysioNet to credentialed researchers who sign the data use agreement. The data cannot be redistributed by the authors. Analysis code and the locked analysis plans are available on GitHub. The frozen model package will be shared through PhysioNet under credentialed access and is available meanwhile from the corresponding author to investigators with MIMIC-IV access.

https://github.com/ping-ai685/stroke-icu-simulator-mimic

https://physionet.org/content/mimic-iv-ed/2.2/

## FUNDING

This research received no specific grant from any funding agency in the public, commercial, or not-for-profit sectors.

## AUTHOR CONTRIBUTIONS (CRediT)

PL conceived the study, curated the data, designed the methods, wrote the analysis software, performed and validated the formal analysis, produced the visualizations, and wrote the original draft. YX contributed to the conception, methodology, and formal analysis. YZ contributed to validation. NR contributed to reviewing and editing the manuscript. YG contributed to validation and revised the manuscript critically for important clinical content in stroke and critical care. All authors reviewed and edited the manuscript, and all authors read and approved the final manuscript.

## CONFLICT OF INTEREST

The authors declare no competing interests.

## ETHICS APPROVAL

The collection of patient information and the creation of the MIMIC-IV database were reviewed by the Institutional Review Board of Beth Israel Deaconess Medical Center, which granted a waiver of informed consent and approved the data sharing initiative. This study is a secondary analysis of de-identified data, for which no further ethics approval or consent was required. The corresponding author completed the required training and signed the PhysioNet data use agreement before accessing the data. The study was not registered.

## PATIENT AND PUBLIC INVOLVEMENT

Patients and the public were not involved in the design, conduct, reporting or dissemination of this study.

## ACKNOWLEDGEMENTS

We thank the School of Public Health, Fudan University, and the Institute of Global Health, University of Geneva, for institutional support.

## Use of artificial intelligence

The authors designed the study and interpreted the results. Claude (Anthropic; Claude Opus 5.5, used through Claude Code) was used to write and edit analysis code, run analyses at the authors’ direction, generate tables and figures, and draft and edit text. For the analysis plans, the authors specified the questions, outcomes, readiness definitions, data split, candidate algorithms and comparators; the tool drafted the written plans, including the selection rule and system checks, before the corresponding results were produced. GPT-6 Pro (OpenAI, through ChatGPT) was used to comment on drafts and to suggest language edits, which the authors reviewed sentence by sentence before accepting them. The analyses ran locally on the controlled-access data, and no individual-level data were sent to either service. The authors reviewed the analysis plans, checked every reported number against the analysis outputs, and reviewed and revised the manuscript in full. The authors take full responsibility for the content. This use will also be declared in the cover letter, as JAMIA requires.

## DATA AVAILABILITY

MIMIC-IV and MIMIC-IV-ED are available through PhysioNet to credentialed researchers who sign the data use agreement (https://physionet.org/content/mimiciv/). The data cannot be redistributed by the authors. Analysis code (Python scripts) and the locked analysis plans will be made available at https://github.com/ping-ai685/stroke-icu-simulator-mimic. The frozen model package (coefficients and model files) contains no patient records but is derived from credentialed data; it will be shared through PhysioNet under credentialed access and is available meanwhile from the corresponding author to investigators with MIMIC-IV access.

S1 Unit list · S2 Readiness definitions (physiological = document version 0.2; stroke-extended = version 0.3; version 0.1 was a draft replaced before any analysis) and literature basis · S3 Both analysis plans with amendment records · S4 Implementation log summary · S5 Module formulas and selection table · S6 System checks · S7 Direct baselines · S8 Ablation table · S9 Parameter uncertainty · S10 Sensitivity analyses 1–3 · S11 Results for 2020 onwards · S12 Post-exit modules · S13 Odds-ratio tables, effect plots and SHAP (Figure S2) · S14 Applicability flags · S15 T3 monitoring intensity by unit and day · S16 TRIPOD+AI, STRESS and RECORD checklists

## Notes

### Competing Interest Statement

The authors have declared no competing interest.

### Clinical Protocols

https://github.com/ping-ai685/stroke-icu-simulator-mimic/blob/main/docs/analysis_plans.md

### Author Declarations

The Institutional Review Board of Beth Israel Deaconess Medical Center (Boston, MA, USA) gave ethical approval for this work (the MIMIC-IV database) and waived the requirement for informed consent. This study is a secondary analysis of de-identified data accessed under the PhysioNet Credentialed Health Data Use Agreement.

