## Supplementary material S3: statistical analysis plans for "Simulation and temporal validation of ICU trajectories after stroke: patient condition, transfer readiness and actual ICU exit"

This file contains English translations of the two statistical analysis plans (originals in Chinese). Part A is the plan for the description of ICU time (version 1.0, locked 6 November 2026) with its amendments. Part B is the plan for the simulator (version 1.0, locked 6 November 2026). Both were locked before any of the corresponding results were produced. References to internal working documents are replaced by a description of their content. "Readiness" refers to transfer readiness; versions 0.2 and 0.3 of its definition correspond to physiological and stroke-extended readiness in the paper (Supplementary S2).

### Part A. Statistical analysis plan for the description of ICU time

#### A0. Status of this plan

- Results obtained during the exploratory data checks were used to settle definitions and feasibility. Analyses run after this plan was locked are the formal analyses and are reported according to this plan.

- Any change after locking is recorded in the revision log with its date, its reason and whether formal results had been seen.
- Some measures computed during exploration (for example the overall T1/T2/T3 shares) are recomputed in the formal analysis. The authors had seen these numbers, and this is stated. Stratified results, the reproducibility test, sensitivity analyses and patient-level measures had not been computed during exploration.

#### A1. Relation to the earlier study of clinical states

- The same stroke ICU cohort is used, and the earlier study is cited.
- The earlier study described dynamic states in the first 72 hours of ICU care; this study describes the composition of time over the whole ICU stay. The questions and outcomes differ.
- The four states are used here only for description (for example, the state distribution when readiness is reached), not as predictors.
- This study observes the whole ICU stay, censored at death; state shares therefore differ from the earlier study because the observation window differs. Drift of background state features after 72 hours is a limitation.

#### A2. Research questions

**Main question:** How much of the ICU time of patients with stroke is explained by observable treatment and monitoring needs? Does the unexplained time follow a stable and reproducible distribution across stroke subtypes, ICU units and phases of the stay?

**Secondary questions:**

1. When do patients first reach readiness (versions 0.2 and 0.3)? How long after reaching it do they leave the ICU?
2. How often is readiness lost again after it is reached, and why?
3. How do the hours of day at which readiness is reached and at which patients leave the ICU differ?

#### A3. Data and study population

| Item | Content |
|---|---|
| Data | MIMIC-IV v3.1, MIMIC-IV-ED 2.2 |
| Cohort | Stroke ICU cohort of the earlier study: adults; ischaemic stroke, intracerebral haemorrhage or subarachnoid haemorrhage (ICD codes, diagnosis position ≤ 3), traumatic intracranial haemorrhage excluded; first ICU stay in the first qualifying admission; ICU stay ≥ 12 hours (n = 6,368) |
| Exclusion | Death recorded at or before ICU admission (no observation time) |
| ICU scope | As defined by MIMIC `icustays`; neuro intermediate and neuro step-down units count as ICU |
| Observation | From ICU admission to the earlier of ICU exit and death, up to 30 days; stays longer than 30 days are truncated at day 30 and counted |

**Main population: all patients**, including those who die in the ICU, so that the analysis does not condition on a future outcome (survival). Results are also reported by ICU outcome (alive exit, ICU death). Measures involving time to actual exit are computed only among alive exits.

#### A4. Definitions

##### A4.1 Time grid

Each patient has one time point every hour on the hour from ICU admission. Conditions are assessed at each time point using only information available up to that point.

##### A4.2 Transfer readiness

As defined in the readiness definition document, version 0.3; the version current at locking is the formal version.

- **Version 0.2:** respiration, circulation (vasopressors), consciousness and metabolism all met.

- **Version 0.3:** version 0.2 plus no continuous intravenous antihypertensive infusion and not within the post-reperfusion monitoring period.
- **Readiness time:** the confirmation time when all conditions have been met continuously for 6 hours.

##### A4.3 Mutually exclusive hourly decomposition (main measure)

Each hour is assigned to exactly one category, according to the conditions at that hour:

| Category | Definition |
|---|---|
| T1, organ-support and related needs | Version 0.2 not met |
| T2, additional observable stroke-specific needs | Version 0.2 met, but continuous intravenous antihypertensive infusion or within the post-reperfusion monitoring period |
| T3, ICU time not explained by observable needs | All version 0.3 conditions met |

T3 is not called "waiting time". The decomposition uses the conditions at each hour, not the 6-hour confirmation rule or the time of first readiness.

##### A4.4 Stratification variables

| Variable | Definition |
|---|---|
| Stroke subtype | Ischaemic stroke / intracerebral haemorrhage / subarachnoid haemorrhage (patients with both intracerebral and subarachnoid haemorrhage counted as intracerebral haemorrhage, n = 26) |
| ICU unit | **The unit in which the hour was spent** (from the `transfers` table), in two types: standard ICU (surgical, neurosurgical, trauma, medical, cardiac vascular, coronary care and similar) and neuro intermediate / step-down. Individual units with ≥ 100 patients are also reported |
| Phase of stay | ICU day 1, days 2–3, 4–7, 8–14, 15–30 |
| Era | MIMIC `anchor_year_group` (2008–2010, 2011–2013, 2014–2016, 2017–2019, 2020–2022) |
| Reperfusion | Thrombolysis or thrombectomy, yes or no (ischaemic stroke only) |

#### A5. Statistical analysis

##### A5.1 Main analysis: composition of T1/T2/T3

Two measures are reported because they answer different questions:

| Measure | Meaning | Computation |
|---|---|---|
| **Hour-based** | Composition of ICU resources (patient-hours) | Share of each category among all hours pooled; longer stays carry more weight |
| **Patient-based** | Composition of a typical patient's ICU time | Each patient's shares, summarised by median, quartiles and mean |

Confidence intervals: patient-level bootstrap, 1,000 resamples, 2.5th and 97.5th percentiles. Results are reported for each stratification variable; unit type and phase of stay are also cross-tabulated.

##### A5.2 Reproducibility across eras

The analysis in A5.1 is repeated in each era group. **"Stable" is defined in advance** as all three of:

1. In every era group, the hour-based T3 share differs from that of the whole cohort by no more than 5 percentage points;
2. The ordering of T3 share between the two unit types is the same in every era group;
3. The direction of change in T3 share across phases of stay is the same in every era group.

If all three hold, the composition is called stable across eras; otherwise the condition that fails is reported. The 2020–2022 group (COVID-19 period) is marked separately.

##### A5.3 Association with patient characteristics (descriptive)

Each patient's T3 share is modelled with a fractional logit model (robust standard errors) on stroke subtype, age, sex, main unit type, reperfusion, era and ICU length of stay. Descriptive only.

##### A5.4 Secondary question 1: timing of readiness

- Cumulative incidence of first reaching versions 0.2 and 0.3, with ICU death as a competing event (Aalen–Johansen), by stroke subtype and unit type.
- Among alive exits, time from first reaching version 0.3 to ICU exit: median, quartiles, and the shares ≤ 6, 6–24 and > 24 hours.
- The time difference between versions 0.2 and 0.3.

##### A5.5 Secondary question 2: loss of readiness

- Hour-based: among hours observed after first reaching version 0.3, the share in which conditions were not met (**a share of hours, not of patients**).
- Patient-based: share of patients with at least one run of ≥ 6 consecutive hours with conditions unmet after first reaching version 0.3.
- Reasons: the conditions unmet when readiness was lost (respiration, vasopressor, antihypertensive infusion, each consciousness component, metabolism), more than one allowed.
- Loss of readiness is not interpreted as deterioration in every case: it may include restarted treatment, fluctuating measurements and missing records.

##### A5.6 Secondary question 3: hour of day

- Distribution over 24 hours of the time of first reaching version 0.3 and of ICU exit.
- Share of exits after 19:00 (the "after hours" definition for 12-hour shifts in the SCCM guideline), by unit. Descriptive only, not related to post-exit outcomes.

#### A6. Sensitivity analyses

| No. | Content | Purpose |
|---|---|---|
| S1 | Hours classed as T1 only because of a missing GCS record classed separately as "insufficient evidence" | Effect of missing data on the T1/T3 shares |
| S2 | Version 0.2 in place of version 0.3 (T1 versus the rest) | Role of the stroke-specific conditions |
| S3 | Reperfusion rule: thrombolysis only; exact administration time where available; no reperfusion rule | Effect of the reperfusion proxy rule |
| S4 | GCS decline threshold ≥ 3 points | Sensitivity of the consciousness condition |
| S5 | Patients with a CPAP order for sleep apnoea: CPAP/BiPAP masks not counted as ICU-level support | Specificity of the respiration condition |
| S6 | GCS validity 4 and 8 hours | Effect of recording interval |
| S7 | Alive exits only | Comparison with the exploratory analysis |
| S8 | Stays truncated at 30 days excluded | Effect of very long stays |
| S9 | Unit classed by last unit rather than hour by hour | Effect of the unit classification |
| S10 | Stable run of 4, 12 and 24 hours (affects only readiness time in secondary question 1, not the hourly decomposition) | Effect of the confirmation rule |

**Methodological note (not a result):** the difference between the hourly decomposition and a split at first readiness is attributed hour by hour to three parts: loss after first readiness; the confirmation time before first readiness; and the handling of unknown values.

#### A7. Rules for interpretation

- T3 is called "ICU time not explained by observable needs", **not waiting time**, and is not interpreted as unnecessary ICU stay; it may contain much necessary neurological monitoring that is not observable.
- No claim is made that patients "could have been transferred earlier".
- No threshold or definition is adjusted according to death, readmission or other outcomes.

- Descriptive and associational analyses only; no causal inference.
- A high T3 share in neuro intermediate units may reflect their function as stroke units and is not interpreted as delay.

#### A8. Reporting

Reporting guideline: RECORD. Planned figures and tables: study flow; baseline characteristics; T1/T2/T3 composition (hour-based and patient-based) by unit type and phase; T1/T2/T3 by each stratification variable with confidence intervals; T3 share by era (reproducibility); cumulative incidence of reaching versions 0.2 and 0.3 (death as competing event); hour-of-day distributions of reaching readiness and of exit; supplementary material with the full readiness definition, sensitivity analyses and item and code lists.

#### A9. Data governance

- MIMIC-IV is controlled-access data: individual-level data are not sent to any third-party AI service, not redistributed and not printed in conversation.
- The AI assistant works only with code, the data dictionary and aggregate results.
- Code for the formal analysis is kept separate from exploratory code; data paths are read from one configuration file.
- Code and aggregate results are made public on publication.

#### A10. Non-analytical items (open at locking, without effect on the analysis)

Title and target journal; the stability threshold in A5.2 (confirmed at locking: 5 percentage points); authors and contributions.

#### A11. Amendment 1.1: separate presentation by unit type (after formal results were seen)

**Reason:** by the pre-specified criteria in A5.2, T3 was **not stable** across eras. A post hoc analysis showed that the main reason was a rise in the share of all hours spent in neuro intermediate / step-down units, from 10–15% before 2017 to 47%, whereas T3 within each unit type was similar across eras. The pooled share therefore depends heavily on the unit mix and is misleading as the only main result.

**Changes:**

1. The main presentation reports T1/T2/T3 **separately by unit type** (standard ICU; neuro intermediate / step-down); pooled results are still reported.
2. The result of the pre-specified test in A5.2 **is not changed**: T3 is reported as not stable across eras.
3. **Post hoc analysis**, labelled as such: conditions 1 and 3 of A5.2 repeated within each unit type, and the share of hours spent in each unit type reported by era.
4. **Hour-based figures are the main figures**, with patient-based figures reported alongside. A5.1 required both without specifying which leads; this is a presentation decision.
5. All other definitions, thresholds and sensitivity analyses are unchanged.

#### A12. Amendment 1.2: two added analyses

**Reason:** the version 1.0 results showed that about half of ICU time was T3, without any link to patient outcomes. Two analyses were added: monitoring intensity within T3, and the clinical meaning of the readiness definition.

**Status when added:** all results of versions 1.0 and 1.1 had been seen; **no result of these analyses had been seen, and readmission and post-exit death data had never been examined in this study.** The readiness definition (version 0.3) and the T1/T2/T3 rules are not changed by these results.

##### A12.1 Monitoring intensity within T3

- For each T3 hour, count the GCS assessments recorded in the preceding 6 hours, including that hour (one per time stamp).

- **High intensity:** ≥ 3 assessments (about every 2 hours or more often). **Usual intensity:** ≤ 2 (about every 4 hours or less).
- Report the shares of high- and usual-intensity T3 hours (hour-based first, patient-based as a supplement), by unit type and phase of stay.
- Sensitivity analysis: ≥ 2 assessments in the preceding 4 hours.
- Interpretation: the number of recorded assessments may differ from the number performed and depends on unit recording practice (hence stratification by unit type). High-intensity T3 is more likely to reflect ICU-level monitoring needs; usual-intensity T3 may include waiting. Neither can be shown directly.

##### A12.2 Clinical meaning of the readiness definition (criterion validity)

- **Population:** patients leaving the ICU alive (stays truncated at 30 days excluded).
- **Exposure:** status at ICU exit, in three groups: (1) reached version 0.3 and still met at the last hour before exit; (2) reached version 0.3 but not met at exit (lost); (3) never reached version 0.3.
- **Outcomes:** primary, ICU readmission in the same hospitalisation (another ICU stay after this one in the same admission); secondary, in-hospital death after ICU exit, and the composite of readmission or death.
- **Analysis:** outcome proportions by group; logistic regression with group 1 as reference, adjusted for age, sex, stroke subtype, log ICU length of stay, unit type at exit, era, reperfusion therapy and exit between 19:00 and 07:00. Odds ratios with 95% confidence intervals. Association only.
- **Secondary exposure:** the analysis repeated with version 0.2.
- **Sensitivity analysis:** patients with a comfort-measures-only code status before ICU exit excluded.
- No definition or threshold is adjusted according to the outcomes.

#### A13. Amendment 1.2.1: data source for the comfort-measures sensitivity analysis

The source of the comfort-measures-only status for the sensitivity analysis in A12.2 was changed from the code-status field of the order table to the Code Status entry of the ICU nursing chart. The order-table field contains only full code, DNR and DNAR, with no comfort-measures option, so the original source identified no patient and the sensitivity analysis had in fact not been run. At this change the main results of A12.2 (readmission, post-exit death) had been seen; the result of the sensitivity analysis with the new source had not.

#### Revision log (Part A)

| Version | Change | Formal results seen |
|---|---|---|
| 0.1 | First draft | No (overall shares from the exploratory analysis had been seen; see A0) |
| 1.0 | Locked; stability threshold confirmed at 5 percentage points | No |
| 1.1 | Main presentation by unit type; pre-specified conclusion of A5.2 unchanged; post hoc within-unit reproducibility; hour-based figures as main figures (A11) | **Yes.** Triggered by the A5.2 result (not stable across eras); a change of presentation after results were seen, with no change to any definition or threshold |
| 1.2 | Monitoring intensity within T3 and criterion validity of readiness added (A12) | Results of 1.0 and 1.1 seen; **no result of the added analyses, and no readmission or post-exit death data, seen** |
| 1.2.1 | Source of comfort-measures-only status corrected (A13) | Main results of A12.2 seen; result of the corrected sensitivity analysis not seen |

### Part B. Statistical analysis plan for the simulator

**Rule after locking:** the plan is amended only for a problem that would cause data leakage, an incorrect outcome definition, model-selection bias or serious leakage of time information, with the effect, reason and version recorded. All other issues go to the implementation log, test cases and the limitations, without changing this plan. (No amendment was made.)

#### B0. Core question and principles

**Core question:** Can explicit modelling of patient state, transfer readiness and care decisions yield a simulation system that is more accurate and more interpretable than conventional ICU length-of-stay prediction and that generates complete patient paths?

**Three levels:** (1) describe the real path; (2) reproduce how the path forms, with disease course and care decisions modelled separately; (3) show whether this design adds value, against direct prediction models and reduced simulators.

**Wording:** "reproducing why a patient leaves the ICU at a given time" means identifying and simulating **observable mechanisms** associated with the timing of exit, not establishing the cause for each patient. The simulator describes and predicts the paths that occur; it does not judge when a patient "should" be transferred.

**Methodological principles:**

1. **The clinical question determines the model structure, not the algorithm.** Each module answers one clinical question (death, condition state, readiness, destination, post-exit outcome).
2. **Simple models and machine learning compete on equal terms**, by criteria written in advance.
3. **Predictive performance, calibration, interpretability and transportability jointly determine model choice.** In an hourly simulation, calibrated probabilities and plausible long-run trajectories matter more than discrimination alone.
4. **Tuning is limited, transparent and reproducible.** No unbounded hyperparameter search and no repeated looks at the temporal validation set. The same plan given to different engineers should yield the same selection process.

**Principle for algorithms, tuning and interpretability:** the structure follows the course of illness, transfer readiness and care decisions of ICU patients with stroke. Candidate algorithms are fixed in advance for each module (regression with splines, penalised regression, gradient-boosted trees). Training, hyperparameter selection and probability calibration are done inside the development set with patient-disjoint splits, judged on probabilistic accuracy, calibration, stability, complexity and the plausibility of complete simulated paths. Where a complex model gives no meaningful and stable gain, the simpler model is preferred. After selection, variable processing, algorithms, hyperparameters, calibration and simulation rules are frozen and evaluated on an independent temporal validation set. External validation uses the same frozen model and pre-defined data mappings; any update or recalibration is reported as a separate analysis. Every model is reported with its clinical meaning, inputs, scope, calibration and main sources of error; machine-learning explanations describe the model's associations, not causal effects.

**Status of this plan:** the authors had seen the results of the path analysis (Part A). **No simulator result (estimated rules, model comparisons, simulations, validation) had been produced.**

#### B1. Clinical use

| Time point | Clinical question | Simulator output |
|---|---|---|
| ICU admission, 24, 48, 72 hours | What will happen to this patient in the next day or two? | Probabilities of **alive exit / ICU death / still in ICU** within 24, 48 and 72 hours |
| Same | How much longer, and where is the patient held up? | Remaining time to the end of the ICU stay, split into time before readiness and time from readiness to exit |
| ICU exit | Will the patient come back soon? | Probabilities of ICU readmission and in-hospital death within 72 hours of exit |
| Daily, unit level (secondary) | How many patients will leave tomorrow? | Expected number of exits among patients in the unit |

Each prediction is accompanied by an explanation of which part (condition state, readiness, hour of day and unit) drives it (B8).

#### B2. Data and split

| Set | Machine-learning equivalent | Content | Patients | Use |
|---|---|---|---|---|
| Development | Training (with internal validation) | Approximate admission year ≤ 2016 | 3,131 | All rules estimated; **algorithm and parameter choice only within this set** |
| Internal training | Training | Approximate admission year ≤ 2014 | 1,877 | **All tuning, early stopping and calibration within each algorithm** (5-fold cross-validation grouped by patient) |
| Internal validation | Validation | Approximate admission year 2015–2016 | 1,254 | **Only the comparison between the three algorithms and the system checks (B4.5); no tuning or calibration.** (ICU deaths 145; ICU readmissions within 72 h of exit 66; in-hospital deaths 42. With fewer than 50 death events, P1b uses nested cross-validation, B4.3) |
| Temporal validation | Test | Approximate admission year 2017–2019 | 1,714 | Final evaluation, **used once** |
| Additional validation | Second test | Approximate admission year ≥ 2020 (COVID-19 period) | 1,521 | Robustness, reported separately |

**Why internal validation is also split by era:** the simulator is meant to predict future patients; an era split mimics this and favours models that remain valid in later years (and in eICU). A random split would place patients from the same era on both sides and make selection optimistic. If a module has fewer than 50 events in the internal validation period, grouped 5-fold cross-validation is used for that module and recorded.

**Approximate admission year** = midpoint of the `anchor_year_group` interval + (shifted admission year − anchor year). Dates in MIMIC are shifted and `anchor_year_group` gives only the 3-year interval of the anchor year, so the precision is about ±1 year; the paper calls it "approximate admission year". Patients near the 2016/2017 boundary may be misclassified; a sensitivity analysis splits by `anchor_year_group` (B10).

##### B2.1 Time conventions (shared by training labels and the simulation engine)

**Hours and rows:** ICU admission is time 0. The k-th hour mark is k hours after admission. Row k is the hour starting at hour mark k; a patient is in the risk set of row k if still in the ICU and alive at hour mark k (end of stay $T > k$).

**Event assignment:** the end of the stay (exit or death) at time T (> 0) is assigned to row $K = \lceil T \rceil - 1$: an event in (k, k+1] belongs to row k. An event exactly at hour mark k+1 belongs to row k, so no event falls in a row from which the patient has already left the risk set.

**Censoring:** stays truncated at 30 days are censored at hour mark 720; their risk set is rows 0–719, without events.

**When the condition state is available:** 6-hour window w covers [6w, 6w+6) and ends at hour mark 6(w+1). Row k uses the filtered state of the last completed window (window $\lfloor k/6 \rfloor - 1$); for $k < 6$ the state is "early admission". The first state is available at hour mark 6 (window 0); transitions between windows start at hour mark 12.

**Fixed order within each row** (training and simulation alike):

| Step | Content | Module | Training label |
|---|---|---|---|
| 1 | k = 6: **first condition state** assigned (state of window 0). k ≥ 12 and divisible by 6: state updated to that of the window just completed | D2-0 (k = 6); D2 (k ≥ 12) | D2-0: state of window 0, patients in the ICU and alive at hour mark 6. D2: transition from window k/6 − 2 to window k/6 − 1, patients in the ICU and alive at hour mark k; first transition at k = 12 |
| 2 | Inputs for the row: state after step 1, version 0.3 status at hour mark k and its duration, readiness confirmed or not, unit, hour of day | — | — |
| 3 | Death in the ICU within the row | D1 | Row k: 1 if death in [k, k+1) |
| 4 | For survivors, destination in the row: stay / move to the other ICU unit type / leave the ICU | C1 | Row k: "leave" if exit in [k, k+1); "move" if the unit type differs between hour marks k and k+1; otherwise "stay". **Rows with a death are excluded from C1 training** |
| 5 | For those staying or moving, version 0.3 status at hour mark k+1, its duration and readiness confirmation updated; movers belong to the new unit from hour mark k+1 | D3 | Row k: patients in the ICU and alive at hour mark k+1; outcome = status at k+1 |
| 6 | Those leaving enter the post-exit phase | P1a, P1b | See below |

**Post-exit timing (P1a, P1b):**

- Time zero $t_0$ = ICU exit. Post-exit row h is $[t_0 + h, t_0 + h + 1)$, h = 0 … 71.
- **P1a:** at exit, either the first subsequent unit is a hospital unit such as a ward (enter P1b), or the patient leaves hospital directly (discharge within 1 hour of ICU exit; not in P1b).
- **P1b event times:** ICU readmission = admission time of the next ICU stay; in-hospital death = time of death; discharge = discharge time. Events are assigned to row **$h = \max(0, \lceil\Delta\rceil - 1)$**, Δ = event time − $t_0$ in hours; events with Δ ≤ 1 (including at $t_0$) are counted in row 0. Observation ends at 72 hours.
- **Several events in one row:** the earliest by exact time; for identical times, death > readmission > discharge.
- Patients with an event recorded before $t_0$ (contradictory records) are excluded from P1b and counted.
- Patients without an event within 72 hours have "none" in rows 0–71.

All modules use only information available at the time; condition states are always the **filtered** states of the frozen model of the earlier study, in training and simulation. Continuous variables such as hours in the ICU enter L1 and L2 as natural splines (4 degrees of freedom) and L3 as raw values; categorical variables use the same one-hot coding in all three.

#### B3. Modules

##### B3.1 D1, death in the ICU

| Item | Content |
|---|---|
| Clinical question | Will this patient die in the ICU in the next hour? |
| Event and time scale | Risk set: patient-hours in the ICU and alive at the hour mark; outcome: death in the next hour; the competing event (exit) is handled by C1; truncation at 30 days is censoring |
| Inputs | Current condition state (including "early admission"), hours in the ICU, age, stroke subtype, current unit type |
| Candidates | L1 logistic; L2 elastic net (interactions: state × hours in ICU, state × unit type); L3 LightGBM binary (age monotone non-decreasing) |
| Grid, internal validation, selection, calibration | B4.2–B4.4 |
| Missing values and scope | All inputs come from the frozen preprocessing, without further missingness; no simulation beyond 720 hours |
| External validation | Frozen; calibration slope, intercept and C statistic reported |

##### B3.2 D2, condition-state transitions

| Item | Content |
|---|---|
| Clinical question | How will the patient's condition state change over the next 6 hours? |
| Event and time scale | As in B2.1, step 1: patients in the ICU and alive at hour mark 6(w+1), moving from the state of window w−1 to that of window w; complete windows only; 4 outcome classes; one model per current state. **D2 estimates transitions conditional on remaining in the ICU, not the unconditional course of illness**; exit and death are handled first, hour by hour, by D1 and C1. D2 probabilities are not interpreted as natural disease progression |
| Inputs | Hours in the ICU, age, stroke subtype, current unit type |
| Candidates | L1 multinomial logistic; L2 elastic-net multinomial (interaction: hours in ICU × unit type); L3 LightGBM multiclass |
| Selection | B4.3 (multiclass log loss); predicted and observed counts reported for rare transitions (observed < 30 times) |
| External validation | Frozen; predicted and observed proportions for each transition |

##### B3.2a D2-0, initial condition state

| Item | Content |
|---|---|
| Clinical question | In which condition state is the patient during the first 6 hours in the ICU? |
| Event and time scale | At hour mark 6; risk set: patients in the ICU and alive at hour mark 6; outcome: filtered state of window 0 (4 classes). Used only when simulating from ICU admission; when simulating from 24, 48 or 72 hours, the patient's actual filtered state is used |
| Inputs | Age, stroke subtype, unit type at admission, reperfusion, source of admission |
| Candidates | L1 multinomial logistic; L2 elastic-net multinomial (no interactions); L3 LightGBM multiclass |
| External validation | Frozen; predicted and observed proportions of the four states |

##### B3.3 D3, version 0.3 readiness status

| Item | Content |
|---|---|
| Clinical question | When does the patient meet the transfer conditions, and are they lost again? |
| Event and time scale | Two sub-models. D3a, risk set: patient-hours with conditions not met; outcome: met in the next hour. D3b, risk set: patient-hours with conditions met; outcome: not met in the next hour |
| Inputs | Current condition state, hours in the ICU, hours in the current status, current unit type, stroke subtype, reperfusion × within 24 hours of ICU admission |
| Candidates | L1 logistic; L2 elastic net (interactions: state × hours in ICU, state × unit type); L3 LightGBM binary |
| Selection | B4.3; D3a and D3b selected separately |
| Missing values | As in the readiness definition (GCS older than 6 hours is "unknown" and counts as not met) |
| External validation | Frozen; availability of each readiness component in eICU audited and reported first, then the pre-defined missing-data rules applied, without ad hoc substitution |

##### B3.4 C1, destination

| Item | Content |
|---|---|
| Clinical question | In this hour, does the patient stay in the current unit, move to the other ICU unit type, or leave the ICU? |
| Event and time scale | Risk set: patient-hours in the ICU and alive; outcome: 3 mutually exclusive classes; ICU death is decided first by D1 (B5) |
| Inputs | Readiness confirmed; time since confirmation (0–6 / 6–24 / 24–72 / > 72 hours); version 0.3 conditions met now; current condition state; hour of day (4-hour blocks); weekend; current unit type; stroke subtype; hours in the ICU |
| Candidates | L1 multinomial logistic; L2 elastic-net multinomial (interactions: readiness × hour block, readiness × unit type, hour block × unit type, state × hours in ICU); L3 LightGBM multiclass |
| Scope | Units mapped to the two types by a pre-specified list; units outside the list flagged as out of |

| Item | Content |
|---|---|
| | scope and reported |
| External validation | Frozen; eICU unit types mapped by a pre-specified table; calibration of exit probability by unit type |

##### B3.5 P1, after ICU exit (evaluated separately from the in-ICU path)

| Item | Content |
|---|---|
| Clinical question | Where does the patient go after the ICU? While still in hospital, will they return to the ICU or die within 72 hours? |
| Event and time scale | **P1a** (once at exit, binary): another hospital unit (ward and similar) / leave hospital directly. Destinations of direct discharges (home, home care, rehabilitation, nursing facility, long-term acute care, hospice, other hospital) are reported descriptively only. **P1b** (patients moving to another hospital unit, hourly after exit): risk set = post-exit patient-hours without an event, still in hospital and within 72 hours; outcome in 4 mutually exclusive classes: **readmission / in-hospital death / discharge / none**, a discrete-time competing-risk multinomial model. **Discharge is a competing event, not "no event"**; events after discharge are not observable |
| Target | Cumulative incidence of readmission and in-hospital death within 72 hours of exit **during the hospitalisation** (discharge as competing event); the full 72-hour risk including after discharge is not estimated |
| Inputs | P1a: condition state at exit, readiness at exit (met / lost / never), log ICU length of stay, stroke subtype, age. P1b: hours since exit, readiness at exit, state at exit, log ICU length of stay, exit between 19:00 and 07:00, unit type at exit, stroke subtype, age, reperfusion |
| Candidates | L1; L2 (P1b interactions: readiness × hours since exit, night exit × readiness); L3 |
| Internal validation | P1a as D1; **P1b, with few events, by nested cross-validation** (B4.3) |
| Scope | Second ICU stays after readmission are not simulated; hospice discharges and transfers to other hospitals are counted separately, and post-exit deaths are interpreted with the possibility of comfort care in mind |
| External validation | Frozen; eICU contains in-hospital information only, so the in-hospital part is evaluated |

Note from the path analysis: 37.7% of patients transferred to a ward were discharged within 72 hours, so discharge is the main competing event. Of 975 patients discharged directly from the ICU, 367 went home, 220 to rehabilitation, 154 to home care, 67 to long-term acute care, 61 to hospice, 53 to a nursing facility, 17 to another acute hospital and 36 elsewhere.

#### B4. Algorithms, tuning, selection and calibration (all modules)

##### B4.1 Three candidates

| Candidate | Algorithm | Why included | Interpretability |
|---|---|---|---|
| **L1** | Logistic / multinomial logistic, splines with 4 degrees of freedom, unpenalised | Rules read as "other things equal, this factor raises or lowers the probability by so much"; external validation needs only intercept and slope updates; individual rules can be edited later | Highest |
| **L2** | Elastic-net logistic with the interactions listed in B3 | Clinical transfer decisions often depend on several factors together; keeps the regression form while letting the data decide whether the interactions are needed | High |
| **L3** | LightGBM with post hoc calibration | Tests for unanticipated non-linearity and interaction | Moderate (SHAP) |

Random forests (tree models like gradient boosting, usually with worse probability calibration) and neural networks as modules (no clear advantage on hourly tabular data with few events, and harder to interpret and transport) are not included. A neural network is kept among the direct prediction models.

#### B4.2 Hyperparameters (all fixed in advance)

**L2:** inputs standardised with the mean and standard deviation of the internal training period; mixing α ∈ {0, 0.5, 1}; penalty λ **fixed at 20 log-spaced values from $10^{-4}$ to $10^{1}$** (not generated by software); solver scikit-learn `saga`, maximum 5,000 iterations, tolerance $10^{-4}$. In 5-fold cross-validation grouped by patient within the internal training period, the **one-standard-error rule** is used: the strongest penalty within one standard error of the minimum cross-validated log loss. Among the three α values the one with the lowest cross-validated log loss under this rule is chosen; ties go to the larger α (sparser).

**L3 (LightGBM):**

| Parameter | Setting |
|---|---|
| num_leaves | **7, 15, 31** (tuned) |
| min_child_samples | **100, 200, 1000** (tuned) |
| learning_rate | 0.05 |
| n_estimators | Up to 1,000, with early stopping on a held-out part **within** the internal training period (B4.4), stopping after 50 rounds without improvement |
| max_depth | −1 (depth controlled by num_leaves) |
| max_bin | 255 |
| feature_fraction | 0.8 |
| bagging_fraction / bagging_freq | 0.8 / 1 |
| min_split_gain | 0 |
| reg_alpha / reg_lambda | 0 / 1 |
| Objective | binary; multiclass |
| Class weights | **None** (original event probabilities kept) |
| Seed, threads | 20261106; deterministic = true; num_threads = 8 |
| Monotone constraint | Only in D1: age → death risk non-decreasing |

Following B4.4, the combination with the lowest **mean cross-validated log loss** among the 9 is selected; ties go to fewer leaves and more samples per leaf (simpler). The number of trees is the median of the 5 early-stopping results.

Patient-hours are not independent; min_child_samples constrains rows, not patients. Internal validation therefore never lets a patient appear on both sides, and patient-level stability is checked in B4.3. Software versions are recorded at run time.

#### B4.3 Module-level selection rule (deterministic)

**Tuning and comparison use different data:** all tuning, early stopping and calibration within each algorithm take place in the internal training period (5-fold cross-validation grouped by patient); the internal validation period is used only once, to compare the three algorithms. The comparison is therefore fair: none of L1, L2 or L3 has used the outcomes of the internal validation period.

For each module (D2-0, D2, D3a, D3b, P1a, P1b separately):

1. Each candidate's parameters and calibrator are set within the internal training period (B4.2, B4.4) and refitted once on the whole internal training period.
2. On the internal validation period compute the **mean log loss** per patient-hour (multiclass log loss where applicable) and the **calibration slope** (binary: slope of the outcome on the predicted log-odds; multiclass: one-versus-rest for each non-reference class, all of which must qualify).
3. **Eligible:** calibration slope between 0.8 and 1.2. This is an **engineering criterion** of this study to exclude clearly over- or under-confident models, not an accepted clinical standard.
4. Among eligible candidates, choose the lowest log loss; **if a more interpretable eligible candidate (order L1 > L2 > L3) is within 1% (relative) of it, choose the more interpretable one.**
5. If none is eligible, use L1 and report the module's calibration problem.
6. **Patient-level stability** (reported, does not change the choice): 95% interval of the log-loss difference between the selected candidate and L1 from 200 patient-level bootstrap samples; calibration by hours in ICU, unit type and risk decile.

**Nested cross-validation for P1b** (few events): outer 5 folds grouped by patient over the whole development set; within each outer training part, inner 5 folds grouped by patient for tuning, early stopping and calibration; evaluation on the outer validation part. Out-of-fold predictions of the 5 outer folds are pooled and steps 2–5 applied. The outcomes of each evaluated fold are never used for tuning or calibration.

**Level of evidence for P1b:** nested cross-validation is used only to choose between the three algorithms; its level of evidence differs from the independent internal validation period used for the other modules, and it does not replace temporal validation. P1b is not part of the A/B system choice (B4.5). After selection, the final P1b model keeps the settings chosen in the outer loop (the most frequent combination in nested cross-validation; ties to the simpler) and is refitted on the whole development set.

#### B4.4 Models with early stopping or calibration: common cross-fitting procedure (L3, direct GBM, direct GRU)

Goal: early stopping, parameter choice and calibration each use data not involved in the corresponding fit; the independent internal validation period is used only for the final comparison of algorithms.

**Round 1, parameters and number of iterations** (within the internal training period):

1. Split the internal training period into 5 folds by patient (seed 20261106).
2. In each fold, hold out 10% of the patients of the training part as an **early-stopping part**; fit on the other 90% and set the number of iterations (trees or epochs) on the early-stopping part.
3. Compute log loss on the validation part of the fold.
4. Choose the candidate setting (9 for L3; 32 or 64 hidden units for the GRU) with the lowest mean log loss across the 5 folds; the number of iterations is the median of the 5 early-stopping results.

**Round 2, calibrator** (within the internal training period, settings fixed):

5. Split the internal training period **again** into 5 folds by patient (seed 20261107).
6. In each fold, fit on the training part with the fixed settings (no early stopping) and predict the validation part.
7. Fit the calibrator on the pooled out-of-fold predictions: Platt scaling for binary outcomes, temperature scaling for multiclass. Every out-of-fold prediction comes from a model that has not seen that patient; the only residual optimism is that these patients took part in the comparison of settings in round 1, which is reported.

**For module selection:** fit once on the whole internal training period with the fixed settings, apply the round-2 calibrator, and evaluate on the independent internal validation period.

**Final model: hyperparameters and number of iterations stay as chosen in round 1 and are not re-selected.** Model parameters are refitted on the whole development set with these settings; the calibrator is refitted on the whole development set as in round 2 (new patient folds, seed 20261107, fixed settings, out-of-fold predictions). Everything is then frozen.

**P1b:** the procedure is run within each outer training part of the nested cross-validation (B4.3).

**L1 and L2:** no post hoc calibration (calibration is only evaluated). L2's α and λ are chosen by 5-fold cross-validation within the internal training period (B4.2); the final model keeps this choice and only refits the coefficients on the whole development set.

If a module selects L3, the frozen package stores the model and its calibrator.

#### B4.5 System-level check: two configurations only

On the internal validation period, two simulators are built from modules fitted on the internal training period and run 1,000 times from each of the four landmarks for every internal-validation patient:

- **Configuration A:** each module with the algorithm chosen in B4.3;
- **Configuration B:** all modules L1.

**Required checks** (for A and B separately):

| Check | Criterion |
|---|---|
| Logic | Zero violations of the checks in B7.3 |
| Valid probabilities | The three mutually exclusive probabilities lie in [0, 1] and sum to 1 |
| Cumulative calibration | Alive exit and ICU death within 72 hours of ICU admission: absolute difference between mean predicted cumulative probability and **observed cumulative incidence** ≤ 0.05 (Aalen–Johansen, alive exit and ICU death as competing events, shorter observation censored) |
| Readiness relapse | Simulated share of patients with at least one run of ≥ 6 consecutive hours with conditions unmet after first confirmation within 10 percentage points of the observed share |
| Length of stay | Simulated median ICU length of stay within 20% of the observed median |

**Decision rule:**

| A | B | Action |
|---|---|---|
| Pass | Pass | Compare the 24-hour alive-exit Brier score pooled over the four landmarks: if A is more than 2% (relative) worse than B, choose B; otherwise choose A |
| Pass | Fail | Choose A |
| Fail | Pass | Choose B |
| Fail | Fail | **Stop; no configuration is declared acceptable.** Return to module diagnostics, amend this plan and complete internal evaluation again before freezing; any change is logged and completed before the temporal validation set is used |

The thresholds (2%, 0.05, 10 percentage points, 20%) are not changed after internal results are seen. P1 is not part of this check; it is selected and evaluated separately (B4.3).

##### B4.6 Parameter uncertainty

The development set is resampled by patient with replacement **200 times**; each time all modules are re-estimated with the **selected algorithms and settings** (no re-tuning; L3 tree numbers and calibrator parameters fixed at those of the final model), and the simulation is rerun.

Two different intervals are reported and named distinctly:

| Interval | Source | Question answered | Use |
|---|---|---|---|
| **Prediction interval** (80%) | 1,000 simulations of the same patient with the final model | Where may this patient's future outcome (for example remaining ICU time) fall? | Uncertainty of remaining time; coverage (B7.2) |
| **Parameter-uncertainty interval** (95%) | 200 bootstrap re-estimations of the rules | Because the rules were estimated from finite data, how wrong may the predicted probability itself be? | Error range of predicted probabilities |

##### B4.7 Frozen model package (saved at the end of this paper, for external validation)

Input variables (dictionary, units, coding, missing-data rules, list and mapping of ICU units); condition states (frozen HMM parameters, preprocessing parameters, state numbering); readiness (all rules of the version 0.3 definition and item numbers); the modules (selected algorithms, parameters, hyperparameters, software versions); calibrators (method and fitted parameters); simulation engine (update order, time step, stopping conditions, random seeds); output (a common format for individual predictions and path summaries).

#### B5. Simulation

- The engine advances hour by hour, **each row strictly in the six-step order of B2.1**: (1) first state at k = 6 (D2-0), update at k ≥ 12 divisible by 6 (D2) → (2) inputs → (3) ICU death (D1) → (4) destination of survivors (C1) → (5) version 0.3 status at the next hour mark for those staying or moving (D3) → (6) post-exit phase for those leaving (P1a; those moving to another hospital unit enter P1b for up to 72 hours, with discharge as a competing event). B2.1 is the only execution specification.

- An internal logic check also compares the result of swapping steps (3) and (4), reported only, without changing the specification.
- Each starting point is simulated **1,000 times**, with fixed random seeds.
- Landmarks: ICU admission, 24, 48 and 72 hours; at 24, 48 and 72 hours only patients still in the ICU and alive, starting from their actual state.

### B6. Comparators and the primary comparison

#### B6.1 Direct prediction models (no paths)

At each landmark, the variables available at that time (age, stroke subtype, condition state, version 0.3 status and its duration, readiness confirmed, unit type, organ support, hours in the ICU, hour of day) are used to predict alive exit and ICU death within 24, 48 and 72 hours.

| Model | Algorithm | Tuning |
|---|---|---|
| Baseline 0 | Empirical length-of-stay distribution by stroke subtype | None |
| Baseline 1-LR | Discrete-time competing-risk multinomial logistic regression | None |
| Baseline 1-GBM | LightGBM with the same 9-setting grid and fixed parameters as B4.2 | Common cross-fitting procedure (B4.4) |
| Baseline 1-GRU | Recurrent neural network: hourly variable vectors from ICU admission to the landmark (same variables as LR, continuous variables standardised on the internal training period), sequence length fixed at 72 hours (front-padded with zeros, with a mask); single GRU layer; output heads for the three classes (alive exit / ICU death / still in ICU) at 24, 48 and 72 hours; loss = sum of the three cross-entropies | Hidden units {32, 64}; dropout 0.2; Adam, learning rate $10^{-3}$, batch 256, up to 50 epochs; hidden units, epochs (early-stopping patience 5) and temperature calibrator set by the common procedure (B4.4) within the internal training period; seed 20261106 |

**Strongest direct model:** after tuning and calibration within the internal training period under the same principles as B4.3, the models are compared on the internal validation period by the log loss of the 24-hour alive-exit probability; the lowest is chosen **before the temporal validation set is used.**

#### B6.2 Primary comparison (single)

**Full simulator (configuration chosen in B4.5) versus the strongest direct model**, on the primary end point (B7.1).

#### B6.3 Secondary comparisons

| Comparison | Question |
|---|---|
| Full simulator vs all-L1 simulator | Does machine learning help the simulator? (not applicable if B was chosen in B4.5) |
| Full vs reduced A (no readiness layer) | Is the readiness layer useful? |
| Full vs reduced B (destination without hour of day, weekend, unit) | Is the care-decision layer useful? |
| Full vs reduced C (readiness never lost after confirmation) | Is recording relapse useful? |
| Among direct models | How much better is machine learning than regression in direct prediction? |

All reduced simulators **re-estimate all modules** with the same selection rule and are evaluated on the same validation set.

### B7. Evaluation

#### B7.1 Primary and key secondary end points

| Type | Measure | Computation |
|---|---|---|
| **Single primary end** | Brier score of the 24-hour **alive-exit** probability | Patient-landmarks of the four landmarks in the temporal validation set pooled; also reported by landmark |

| Type | Measure | Computation |
|---|---|---|
| point | | |
| Key secondary end point | Wasserstein distance between simulated and observed ICU length-of-stay distributions (hours) | Simulated distribution from ICU admission against the observed distribution of the temporal validation set; quantiles also reported. For direct models, the distribution is derived from their cumulative predicted risks |

The primary comparison is concluded on the primary end point only; the key secondary end point describes how well paths are reproduced and does not change the primary conclusion.

**"Better":** the difference between two approaches on the same validation set, with a 95% confidence interval from 1,000 patient-level bootstrap samples; only an interval excluding 0 is called better or worse.

**Calibration reference** (descriptive): slope 0.8–1.2, absolute intercept ≤ 0.05.

##### B7.2 Secondary measures

1. Calibration curves and Brier scores of the three mutually exclusive probabilities (alive exit, ICU death, still in ICU) at 24, 48 and 72 hours; C statistics for alive exit and ICU death.
2. Remaining time to the end of the ICU stay: mean absolute error of the predicted median (by way of ending); coverage of the 80% prediction interval (reference 75–85%).
3. Population-level reproduction: state shares at each landmark; cumulative incidence of first readiness; share losing readiness; T1/T2/T3 composition; hour of day of exit; share and timing of ICU death; patient-hours by unit type; readmission and in-hospital death within 72 hours of exit.
4. Predictions at exit (P1): calibration and discrimination for readmission and in-hospital death within 72 hours.
5. Unit level: expected and actual daily numbers of exits.
6. All of the above by unit type (standard ICU / intermediate) and stroke subtype.

##### B7.3 Internal logic checks (all must be zero)

Events after death; readiness confirmed without 6 continuous hours; in-ICU state updates after exit; post-exit phase longer than 72 hours; three probabilities not summing to 1. The effect of deciding death before or after destination on the exit probability is also compared.

#### B8. Interpretability outputs

| Level | Output |
|---|---|
| Rules | L1/L2 modules: odds-ratio tables and effect plots (for example, the probability of leaving the ICU by hour of day once readiness is reached). L3 modules (if selected): SHAP summary and dependence plots for the main variables, compared with the direction in L1; directions contrary to clinical expectation listed and discussed |
| Individual | Remaining time split into before readiness and from readiness to exit, showing where the patient is held up; simulated paths of 2–3 typical patients (simulation results and summaries only, no personal data) |
| System | What each module does, how patients move between states and units, why readiness is lost again; the reduced-simulator comparisons (B6.3) show what each layer contributes |
| Scope | Each prediction flagged when outside reliable use: unit not in the list, more than 720 hours in the ICU, inputs missing beyond the rules, landmark other than the four pre-specified ones |

#### B9. External validation (specified in advance for the next paper)

- The frozen model package (B4.7) is run first, and external validation without any update is reported.
- Only variable harmonisation and pre-defined mappings are allowed (the harmonisation dictionary and frozen preprocessing of the earlier eICU study); no re-tuning, no re-selection of algorithms, no changes in response to eICU results.
- When an input is unavailable in eICU, coverage is reported and the pre-defined missing-data rules applied, without substituting variables or refitting to make the model run.
- The only pre-specified update is recalibration of each module's intercept and slope, reported as a clearly labelled model-updating experiment separate from the frozen-model results.

#### B10. Sensitivity analyses

1. Split by `anchor_year_group` (2008–2016 / 2017–2019).
2. Version 0.2 in place of version 0.3 for D3.
3. Half the number of parameter-uncertainty resamples (100), to check the stability of the intervals.

#### B11. Rules for interpretation

- Only "observable mechanisms associated with the timing of exit" are claimed, not causes.
- No recommendation on when patients should be transferred is derived from the simulations.
- The simulator does not reproduce individual treatment trajectories; second ICU stays after readmission are not simulated.
- If the full simulator is not better than the strongest direct model on the primary comparison, this is reported as it stands, and its value for interpretation and for generating complete paths is discussed.
- If a module selects L3, SHAP and similar explanations are supportive only and are not interpreted as causal effects.

#### B12. Computing resources

All analyses run on a local workstation (18 cores, 48 GB memory) without GPU or cloud computing. The most demanding step is the 200 parameter-uncertainty resamples, run in parallel overnight.

#### B13. Reporting guidelines

TRIPOD+AI (prediction); STRESS (simulation); RECORD (routinely collected data).

#### Revision log (Part B; all versions dated 6 November 2026, before any simulator result)

| Version | Change |
|---|---|
| 0.1 | First draft |
| 0.2 | Clinical use cases; three candidate algorithms per module with reasons; fixed small grids and selection rule (era-based internal validation, log loss, calibration slope, 1% rule favouring interpretability); direct models extended to logistic regression, LightGBM and GRU; primary comparison = simulator versus strongest direct model; interpretability outputs; external validation with intercept and slope recalibration only; computing resources |
| 0.3 | Four methodological principles; module specifications with ten items each (clinical question, event and time scale, inputs, candidates, grid, internal validation, selection, calibration, missing values and scope, external validation); P1 split into P1a and P1b and evaluated separately; LightGBM grid of 9 settings with all fixed parameters; deterministic selection rule and patient-level stability check; leakage-free L3 calibration; system level limited to A versus B with required checks and the 2% rule; frozen model package; applicability flags |
| 0.3.1 | All tuning, early stopping and calibration moved inside the internal training period, internal validation used only for comparison (correcting reuse of internal validation for L2/L3 tuning and L3 calibration); nested cross-validation for P1b; discharge added as a competing event in P1b; P1a redefined; D2 stated as conditional on remaining in the ICU; complete A/B decision table with stop rule; fixed λ grid and solver, LightGBM threads and all GRU settings; calibration slope labelled an engineering criterion; Aalen–Johansen for observed 72-hour cumulative incidence; single primary end point and key secondary end point; prediction and parameter-uncertainty intervals distinguished; wording on approximate admission year |
| 0.3.2 | Time conventions (B2.1) fixing hour marks, rows, state availability and the six-step order within each row, and the risk set of every training label; P1b timing rules; common two-round cross-fitting procedure for L3, GBM and GRU |
| 0.3.3 | Technical corrections: execution order unified with B2.1; first state assignment at hour 6 (new module D2-0) separated from window transitions from hour 12; event assignment $K = \lceil T \rceil - 1$ and censoring rows 0–719; final models keep the hyperparameters and iterations chosen in the internal training period; level of evidence of P1b nested cross-validation |
| 1.0 | Locked (content of 0.3.3) with the rule for changes after locking |
