## Supplemental Data 1 for "Simulation and temporal validation of ICU trajectories after stroke: patient condition, transfer readiness and actual ICU exit"

Abbreviations: AIS, acute ischaemic stroke; GBM, gradient boosting machine; GRU, gated recurrent unit; ICH, intracerebral haemorrhage; L1, logistic regression with natural cubic splines; L2, elastic net; L3, LightGBM; LR, logistic regression; SAH, subarachnoid haemorrhage.

---

#### S1. Units counted as ICU time

ICU time was the time spent in any of the units below (MIMIC-IV `transfers.careunit`). Neuro Intermediate and Neuro Stepdown form the "neuro intermediate / step-down" unit type; all others form the "standard ICU" type.

**Table S1. Units counted as ICU time, with the number of stays that spent any time in each unit and the hourly decomposition of ICU time (units with ≥100 stays).**

| Unit | Type | Stays | Patient-hours | T1, % of hours | T2, % of hours | T3, % of hours (95% CI) |
|---|---|---|---|---|---|---|
| Surgical Intensive Care Unit (SICU) | Standard | 2,250 | 256,887 | 58.0 | 5.3 | 36.7 (34.5–38.6) |
| Neuro Intermediate | Intermediate | 1,846 | 200,481 | 8.4 | 3.2 | 88.4 (87.4–89.3) |
| Neuro Surgical Intensive Care Unit (Neuro SICU) | Standard | 1,521 | 184,295 | 61.2 | 4.5 | 34.3 (32.2–36.5) |
| Trauma SICU (TSICU) | Standard | 703 | 52,084 | 60.4 | 6.4 | 33.2 (30.0–36.8) |
| Medical Intensive Care Unit (MICU) | Standard | 460 | 43,555 | 73.7 | 1.9 | 24.4 (21.1–27.6) |
| Neuro Stepdown | Intermediate | 418 | 34,655 | 12.2 | 4.7 | 83.1 (80.8–85.2) |
| Cardiac Vascular Intensive Care Unit (CVICU) | Standard | 287 | 40,438 | 69.3 | 2.1 | 28.5 (25.2–32.0) |
| Coronary Care Unit (CCU) | Standard | 230 | 19,024 | 62.4 | 1.2 | 36.3 (31.2–42.2) |

| Unit | Type | Stays | Patient-hours | T1, % of hours | T2, % of hours | T3, % of hours (95% CI) |
|---|---|---|---|---|---|---|
| Medical/Surgical Intensive Care Unit (MICU/SICU) | Standard | 199 | 15,205 | 67.7 | 0.5 | 31.7 (26.9–37.6) |

The unit "Intensive Care Unit (ICU)" was also included (standard type) but had fewer than 100 stays. A stay can contribute to more than one unit. 95% CIs from 1,000 patient-level bootstrap samples.

---

### S2. Readiness definitions and literature basis

#### Basis

The structure follows the Society of Critical Care Medicine guideline: a patient can leave the ICU when they no longer meet ICU admission criteria and meet the criteria for a lower level of care.[1] The four physiological conditions follow the operational readiness criteria of the OPENS-2 trial protocol.[14] The criterion for reperfusion therapy follows the AHA/ASA 2019 guideline, which recommends at least 24 hours of monitoring in an ICU or stroke unit after intravenous thrombolysis;[15] it is used here as a conservative proxy for transfer to a general ward, not as a statement that every patient must stay 24 hours in an ICU. Thresholds and durations are design choices of this study, not validated transfer cut-offs.

#### Physiological readiness (definition document version 0.2)

All four conditions had to hold at the same time. Each hour on the hour, only information recorded up to that hour was used.

| Condition | Not met when |
|---|---|
| Respiration | Invasive ventilation in progress; or, in the previous 6 hours, an endotracheal tube, high-flow nasal cannula, BiPAP or CPAP mask, or any ventilator mode (other than standby) recorded |
| Circulation | Continuous infusion of a vasoactive drug in progress (norepinephrine, epinephrine, phenylephrine, vasopressin, dopamine, dobutamine or milrinone) |
| Consciousness | No GCS recorded in the previous 6 hours (status unknown); or a confirmed new GCS decline in the previous 6 hours; or a continuous sedative infusion in progress (propofol, midazolam, lorazepam, dexmedetomidine); or RASS ≥ +3 in the previous 6 hours; or a seizure recorded in the previous 24 hours |
| Metabolism and blood | Most recent result in the critical range (sodium < 125 or > 155 mmol/L; potassium < 3.0 or > 6.0 mmol/L; glucose < 54 or > 400 mg/dL; arterial pH < 7.25 or > 7.55; lactate ≥ 4 mmol/L; haemoglobin < 7 g/dL; platelets < 50 × $10^9$/L); or hypertonic saline, mannitol or a blood transfusion in the previous 6 hours; or a continuous intravenous insulin infusion in progress |

GCS decline: a fall of 2 or more points from the median of the previous 12 hours (at least two records), using the total score when the verbal score could be assessed and eye plus motor scores otherwise (for example, when intubated). The decline was confirmed when the next assessment within 6 hours was still lower; the confirmation time was the time of that reassessment.

Validity of measurements: a GCS record was valid for 6 hours. Electrolytes and blood counts were valid for 24 hours. Blood gases and lactate were assessed only when measured; a missing test did not prevent readiness. A critical result did not revert to normal when it expired, only when a later normal result was recorded. When several results were recorded at the same time, the most abnormal was used.

#### Stroke-extended readiness (definition document version 0.3)

Physiological readiness, plus two stroke-specific conditions:

| Condition | Not met when |
|---|---|
| Blood pressure | Continuous intravenous antihypertensive infusion in progress (nicardipine, clevidipine, labetalol, esmolol or nitroprusside); single intravenous boluses and diltiazem infusions (used mainly for rate control) did not count |
| Reperfusion | Within the first 24 hours after ICU admission for patients with reperfusion therapy whose timing could be placed (evidence grade A or B below); if the procedure date fell after ICU admission, the 24 hours started at 00:00 of that date |

Evidence grades for the timing of reperfusion: A, a reliable administration time (thrombolytic dose > 4 mg); B, a procedure date from the ICD procedure code between the day before and 3 days after ICU admission, or thrombolysis at another hospital with ICU admission within 24 hours of hospital admission; C, treatment known but timing not placeable. The 24-hour rule was applied to grades A and B only.

##### Confirmation

Readiness was confirmed when all conditions had been met at every hour of a continuous 6-hour run; the confirmation time was the sixth hour. After confirmation, readiness was counted as lost in any hour in which a condition was not met; the paper reports losses lasting 6 or more consecutive hours.

##### Revision history of the definition

| Version | Change | Reason | Outcome data seen |
|---|---|---|---|
| 0.1 | First draft | — | No |
| 0.2 | Common end point; two-layer structure; validity rules; non-invasive ventilation and high-flow oxygen added | Discussion and data audit | No |
| 0.3 | Continuous intravenous antihypertensive infusion and the post-reperfusion window added; classification of missing GCS corrected | The distribution of version 0.2 readiness was implausible (about half of patients ready within 7 hours of ICU admission; 99.6% ready in neuro intermediate units) | No: revised after seeing the distribution of readiness itself, not of deaths, readmissions or other outcomes |

Version 0.2 also described a second, "neuro-intervention" layer (active external ventricular drainage or intracranial pressure monitoring in the previous 12 hours). It was computed descriptively but is not part of either definition used in this paper. The neuro-monitoring frequency ordered by clinicians is not recorded in MIMIC-IV and could not be included.

---

#### S3. Statistical analysis plans and amendments

The full English translations of both plans are provided as a separate supplementary file (Supplementary material S3: statistical analysis plans). Both were locked on 6 November 2026, before any of the corresponding results were produced.

**Table S3. Amendments after locking.**

| Plan | Amendment | Change | Results seen at the time |
|---|---|---|---|
| Description of ICU time | 1.1 | Main presentation separated by unit type; pre-specified reproducibility result unchanged ("not stable across eras"); post hoc within-unit reproducibility added; hour-based figures as main figures | Yes: triggered by the reproducibility result. A change of presentation only; no definition or threshold changed |
| Description of ICU time | 1.2 | Two analyses added: monitoring intensity within T3 hours (S15) and criterion validity of readiness at exit (Table 3) | Earlier results seen; results of the added analyses, and all readmission and post-exit death data, not yet seen |
| Description of | 1.2.1 | Source of comfort-measures-only status for | Main criterion-validity results |

| Plan | Amendment | Change | Results seen at the time |
|---|---|---|---|
| ICU time | | the sensitivity analysis of criterion validity corrected (the order-table field has no comfort-measures option, so the original source identified no patient) | seen; result of the corrected sensitivity analysis not seen |
| Simulator | — | None | — |

#### S4. Implementation log summary

The analysis plan was not changed after it was locked. The implementation log records engineering choices that the plan left open, tests, and data features found along the way. The plan allowed amendment only for data leakage, an incorrect outcome definition, selection bias or serious leakage of time information; none of the items below met these conditions.

**Data features recorded before validation.** The data audit (label-consistency checks; 26 of 26 passed after the two checks added during the end-to-end test) showed that care structure changed between eras. Moves between unit types within an ICU stay numbered 3 in the internal training period (up to 2014), 121 in 2015–2016, 618 in 2017–2019 and 642 from 2020; direct hospital discharge from an ICU-level unit rose from 5% and 7% of alive exits to 31% and 26%. Before any validation result was seen, the log recorded the expected consequences: the destination (C1) and direct-discharge (P1a) modules would be hard to estimate and likely miscalibrated in later years, and results would be reported by unit type to separate a change in care structure from a failure of the rules.

**Tests and corrections.**

- End-to-end test (internal training data only): the condition-state transition at hour 720 of stays truncated at 30 days had entered the training table, although the risk set ends at hour 719. It was removed and two audit checks were added. Module fitting was set to fail on missing values rather than drop rows silently.
- Engine speed-up: L1 modules were evaluated through precomputed tables of each term's contribution. A missing constant column was found and corrected; afterwards module probabilities matched the original engine to within $3 \times 10^{-15}$, and 2,000 simulated paths with the same random seed were identical.
- Two programming errors in the direct models (a duplicated column and a sequence index) were corrected without changing the method.
- Refits of the two elastic-net modules differed between runs by up to $3 \times 10^{-7}$ in the coefficients (randomised solver); the frozen coefficients were used throughout.

**Choices the plan left open.**

| Item | Implementation |
|---|---|
| L2 penalty | λ as in glmnet (mean loss + λ × penalty), converted to scikit-learn as $C = 1/(n \times \lambda)$; 20 log-spaced values from $10^{-4}$ to $10^{1}$ |
| L2 standardisation | Within each fold, mean and standard deviation of the training part |
| Folds | Grouped by patient; patients randomly ordered (seed 20261106; second round 20261107) and assigned by position modulo 5 |
| L3 early stopping | 10% of patients held out within each training part |
| L3 inputs | Continuous time variables as raw values; categorical variables with the same treatment coding as L1 |
| Multiclass calibration slope | Reference class = most frequent class in the internal training period; for each other class, logistic regression of the class indicator on logit(p) |
| Classes absent from training | Predicted probability set to 0 (truncated at $10^{-15}$) when computing log loss |
| Simulation from 24, 48 or 72 hours | The observed state at the starting hour was used; the engine skipped the state update in that hour |
| Direct LR and GBM | Discrete-time competing-risk form: hourly predictions of stay, alive exit or ICU death from the landmark to 71 hours, covariates fixed at the landmark plus hours elapsed |

| Item | Implementation |
|---|---|
| Direct GRU | Hourly inputs for up to 72 hours before the landmark (zero-padded and masked), with age, subtype and landmark as static inputs; three output heads for 24, 48 and 72 hours |
| Length-of-stay distance | Direct models predict only to 72 hours, so the Wasserstein distance was also reported up to 72 hours to allow comparison |
| Parameter uncertainty | 300 temporal-validation patient-landmarks (75 per landmark, fixed seed), because all 5,543 would have taken more than 100 hours |
| Sensitivity analyses 1 and 2 | The validation script compared the simulator with GBM by default; as the plan requires comparison with the strongest direct model, the comparison with the GRU was recomputed from the saved predictions with the same bootstrap and seed |

**Observations recorded before temporal validation.** On the internal validation period the simulator's 24-hour Brier score (0.1261) was about 4% worse than that of GBM (0.1210). The log noted that the primary comparison might therefore not favour the simulator and that such a result would be reported as it stood. In two condition-state modules, L2 was selected at the strongest penalty in the grid (λ = 10), close to predicting the base rates; this boundary value is reported.

**Post hoc explanatory calculation (not pre-specified; not used for any conclusion).** To interpret the size of the ablation differences, the 24-hour Brier score was compared with a prediction that gives every patient the development-period exit rate (19.1%; Brier 0.1266; observed rate 14.6%). After a post hoc shift of each model's overall level to the observed rate, the improvement over a single common probability was 6.7% for the full simulator, 4.1% without readiness inputs, 6.2% without clock, weekend and unit, 6.0% without readiness relapse, and 8.1% for GBM. Removing readiness mainly reduced discrimination; removing clock, weekend and unit mainly worsened the overall level.

**Pre-specified items run after co-author review.** While this supplement was prepared, four secondary items specified in the simulator plan were found not to have been run: baseline 0, the swap of the death and destination steps, daily expected exits per unit, and the report of rare state transitions. At the authors' decision they were run on 17 November 2026 with the frozen models, without any other change (S5, S6, S7). Daily exits per unit could not be computed as specified because of per-patient date shifting; the closest feasible quantity is reported.

**Patient-hour count.** The hourly decomposition of ICU time used rows 0 to $\lfloor T \rfloor$ of each stay, the simulator rows 0 to $\lceil T \rceil - 1$. They differ by one boundary row in 86 stays (72 truncated at 720 hours, 14 ending exactly on the hour), giving 847,469 and 847,383 rows. The simulator's count is correct; the difference is 0.01% of hours and changes no proportion by more than 0.01 percentage points.

**Correction during the preparation of this supplement.** The log had summarised the agreement between SHAP and logistic contributions as "Spearman 0.7–0.98". The table (S13h) shows 0.18–0.98 for all variables (0.55–0.98 for the leading variable of each module), with one exception (−0.08). The direction finding is unchanged; the manuscript reports the correct ranges.

---

### S5. Module formulas and algorithm selection

#### Module formulas (L1)

State codes used in the tables: 0, neurological impairment with renal dysfunction; 1, neurological impairment with respiratory support; 2, neurological impairment with low support; 3, neurologically preserved with low support; −1, before the first state is assigned at hour 6.

All continuous time variables enter as natural cubic splines with 4 degrees of freedom in L1 and L2, and as raw values in L3. Categorical variables use treatment coding. Variable names: *state*, current condition state (−1 before hour 6, then 0–3); *k*, hours since ICU admission; *inter*, currently in a neuro intermediate / step-down unit; *run*, hours in the current readiness run (met or unmet); *ever*, readiness ever confirmed; *since*, time since first confirmation (none, 0–6, 6–24, 24–72, ≥72 h); *met*, readiness currently met; *clock*, hour of day in 4-hour blocks; *weekend*; *age* (per 10 years); *subtype* (AIS, ICH, SAH); *reperf*, reperfusion therapy; *reperf<24h*, within 24 hours of reperfusion; *entry*, source of ICU admission.

| Module | Outcome | Formula (L1) |
|---|---|---|
| D1 | Death in the next hour | state + spline(k) + age + subtype + inter |
| D2-0 | Condition state at hour 6 (4 classes) | age + subtype + inter + reperf + entry |
| D2 (one module per current state) | Next condition state, every 6 h (4 classes) | spline(k) + age + subtype + inter |
| D3a / D3b | Readiness gained / lost in the next hour | state + spline(k) + spline(run) + inter + subtype + reperf<24h |
| C1 | Destination: stay, move to the other unit type, leave the ICU | ever + since + met + state + clock + weekend + inter + subtype + spline(k) |
| P1a | Direct hospital discharge within 1 h of ICU exit | state at exit + readiness at exit (met / lost / never) + log ICU length of stay + subtype + age |
| P1b | Hourly outcome for 72 h after exit: none, ICU readmission, in-hospital death, discharge | spline(h since exit) + readiness at exit + state at exit + log ICU length of stay + exit 19:00–07:00 + exit from intermediate unit + subtype + age + reperf |

L2 added pre-listed interactions: D1, state × spline(k) and state × inter; D2, spline(k) × inter; D3, state × spline(k) and state × inter; C1, ever × clock, ever × inter, clock × inter and state × spline(k); P1b, readiness at exit × spline(h) and exit 19:00–07:00 × readiness at exit.

#### Algorithm selection

Candidates were tuned and calibrated on the internal training period (approximate admission year up to 2014) and compared once on the internal validation period (2015–2016); P1b used nested cross-validation on the whole development set. A candidate was eligible if every calibration slope lay between 0.8 and 1.2. Among eligible candidates the one with the lowest log loss was chosen, unless a more interpretable one was within 1%; if none was eligible, L1 was used.

**Table S5. Candidate algorithms by module: validation log loss, calibration slopes (by outcome class), eligibility and selection.**

| Module | Candidate | Log loss | Calibration slope(s) | Eligible | Selected | Reason / settings |
|---|---|---|---|---|---|---|
| D1 | L1 | 0.0063 | 1.02 | Yes | ✓ | L3 lowest, L1 within 1% |
| D1 | L2 | 0.00649 | 1.03 | Yes | | α=1.0, λ=3.36e-04 |
| D1 | L3 | 0.00624 | 1.02 | Yes | | leaves 7, min samples 200, 103 trees, Platt |
| D2-0 | L1 | 1.05869 | 0.83; 0.88; 0.84 | Yes | ✓ | lowest eligible |
| D2-0 | L2 | 1.06871 | 1.69; 1.62; 1.80 | No | | α=0.5, λ=4.28e-02 |
| D2-0 | L3 | 1.06154 | 1.30; 0.98; 1.23 | No | | leaves 15, min samples 100, 25 trees |
| D2, from 0 (renal dysfunction) | L1 | 0.68855 | 0.64; 0.27; 0.03 | No | | |
| D2, from 0 (renal dysfunction) | L2 | 0.68712 | 0.88; 0.93; 0.86 | Yes | ✓ | lowest eligible; α=0.0, λ=1.00e+01 |
| D2, from 0 (renal dysfunction) | L3 | 0.68645 | 0.60; 0.92; 0.26 | No | | leaves 7, min samples 200, 18 trees |
| D2, from 1 (respiratory support) | L1 | 0.64323 | 0.89; 0.58; 0.72 | No | | |
| D2, from 1 (respiratory | L2 | 0.65232 | 2.20; 0.94; 8.52 | No | | α=1.0, λ=4.28e-02 |

| Module | Candidate | Log loss | Calibration slope(s) | Eligible | Selected | Reason / settings |
|---|---|---|---|---|---|---|
| support) | | | | | | |
| D2, from 1 (respiratory support) | L3 | 0.6442 | 1.01; 0.93; 0.89 | Yes | ✓ | lowest eligible; leaves 7, min samples 1000, 55 trees |
| D2, from 2 (impairment, low support) | L1 | 0.86378 | 0.68; 1.31; 0.17 | No | ✓ | none eligible, L1 by rule |
| D2, from 2 (impairment, low support) | L2 | 0.87762 | 60.55; 18.49; −0.29 | No | | α=0.0, λ=1.00e+01 |
| D2, from 2 (impairment, low support) | L3 | 0.83911 | 0.94; 0.97; 0.77 | No | | leaves 7, min samples 1000, 1 tree |
| D2, from 3 (preserved) | L1 | 0.34104 | 1.03; 0.95; 0.60 | No | | |
| D2, from 3 (preserved) | L2 | 0.34937 | 0.95; 0.95; 1.00 | Yes | ✓ | lowest eligible; α=0.0, λ=1.00e+01 |
| D2, from 3 (preserved) | L3 | 0.34266 | 1.07; 0.32; 0.50 | No | | leaves 7, min samples 1000, 64 trees |
| D3a (gained) | L1 | 0.1009 | 0.96 | Yes | | |
| D3a (gained) | L2 | 0.1022 | 1.45 | No | | α=0.0, λ=1.27e-02 |
| D3a (gained) | L3 | 0.08803 | 1.01 | Yes | ✓ | lowest eligible; leaves 15, min samples 100, 134 trees, Platt |
| D3b (lost) | L1 | 0.12509 | 1.11 | Yes | | |
| D3b (lost) | L2 | 0.12653 | 1.80 | No | | α=0.5, λ=6.95e-03 |
| D3b (lost) | L3 | 0.12283 | 1.15 | Yes | ✓ | lowest eligible; leaves 7, min samples 1000, 153 trees, Platt |
| C1 | L1 | 0.04304 | 0.60; 0.99 | No | ✓ | none eligible, L1 by rule |
| C1 | L2 | 0.04227 | 0.77; 0.92 | No | | α=0.0, λ=3.79e-03 |
| C1 | L3 | 0.04277 | 0.55; 1.04 | No | | leaves 7, min samples 1000, 137 trees |
| P1a | L1 | 0.25964 | 0.38 | No | ✓ | none eligible, L1 by rule |
| P1a | L2 | 0.23572 | 1.28 | No | | α=1.0, λ=2.34e-02 |
| P1a | L3 | 0.24719 | 0.50 | No | | leaves 15, min samples 100, 52 trees, Platt |
| P1b | L1 | 0.04607 | 0.98; 0.99; 0.96 | Yes | | |
| P1b | L2 | 0.04687 | 0.98; 1.00; 1.50 | No | | most frequent: α=1.0, λ=6.16e-04 |
| P1b | L3 | 0.04335 | 0.97; 0.98; 1.11 | Yes | ✓ | lowest eligible; most frequent: leaves 15, min samples 1000, 72 trees |

All 16 condition-state transitions occurred at least 30 times in the development set (57,839 transitions), so the planned report of rare transitions (fewer than 30) was empty. Calibration slopes for multiclass modules are given for each non-reference class. L3 calibration used Platt scaling (binary) or temperature scaling (multiclass), fitted on cross-fitted predictions.

---

#### S6. System checks

Both configurations were fitted on the internal training period and simulated 1,000 times from 0, 24, 48 and 72 hours for every patient in the internal validation period (3,737 patient-landmarks). Configuration A would be retained unless its Brier score was more than 2% (relative) worse than that of configuration B.

**Table S6. System checks on the internal validation period.**

| Measure | Configuration A (per-module choice) | Configuration B (all L1) |
|---|---|---|
| 24-h alive-exit Brier score (four landmarks pooled) | 0.1261 | 0.1260 |
| 72-h alive exit, simulated / observed | 0.448 / 0.469 | 0.464 / 0.469 |
| 72-h ICU death, simulated / observed | 0.080 / 0.061 | 0.076 / 0.061 |
| Readiness relapse among ever ready, simulated / observed | 0.481 / 0.394 | 0.472 / 0.394 |
| Median ICU length of stay (h), simulated / observed | 69 / 67 | 67 / 67 |
| Logical violations in simulated paths | 0 | 0 |

**Order of death and destination within each hour.** The plan specified a check in which the death step and the destination step are swapped. It was run after the main analysis (17 November 2026) on the temporal validation set with the same frozen model and random seeds. Mean predicted probabilities changed by at most 0.0003 (24-h alive exit 0.2224 vs 0.2227; 72-h ICU death 0.0489 vs 0.0488), the mean absolute difference per patient-landmark was 0.013–0.015, and the 24-h alive-exit Brier score was 0.1238 vs 0.1237.

Required checks (both configurations passed all): no logical violation; valid probabilities; 72-hour cumulative calibration for alive exit and ICU death within 0.05; readiness relapse within 10 percentage points; median length of stay within 20%. Configuration A was retained.

---

#### S7. Direct prediction models

Three direct models predicted the same outcomes at each landmark without generating paths: logistic regression and gradient boosting in discrete-time competing-risk form, and a gated recurrent unit network using the preceding 72 hours. They were tuned on the internal training period and compared on the internal validation period (3,737 patient-landmarks). The model with the lowest log loss for the 24-hour alive-exit probability was the comparator for the primary analysis (GBM). Settings: GBM 31 leaves, minimum 1,000 samples, 142 trees; GRU 64 hidden units, 28 epochs, temperature 1.013.

**Table S7. Direct models on the internal validation period.**

| Model | 24 h log loss | 24 h Brier | 24 h slope | 48 h log loss | 48 h Brier | 48 h slope | 72 h log loss | 72 h Brier | 72 h slope |
|---|---|---|---|---|---|---|---|---|---|
| LR | 0.39514 | 0.12526 | 1.31 | 0.53302 | 0.18 | 1.01 | 0.55434 | 0.18674 | 0.94 |
| GBM | 0.38046 | 0.12103 | 0.98 | 0.5377 | 0.18106 | 0.86 | 0.56329 | 0.18896 | 0.85 |
| GRU | 0.39579 | 0.12391 | 0.89 | 0.52606 | 0.17818 | 1.01 | 0.54132 | 0.18259 | 1.01 |

**Baseline 0 (empirical exit rates).** The plan also specified a baseline without patient-level information: each patient-landmark receives the share of development patient-landmarks with the same stroke subtype and landmark that left the ICU alive, or died, within 24, 48 and 72 hours. It was run after the main analysis (17 November 2026, see S4). On the internal validation period its 24-h alive-exit log loss was 0.45395 (Brier 0.14363), so GBM remained the strongest direct model and the primary comparison was unaffected.

**Table S7b. Baseline 0 in the validation sets.**

| Measure | 2017–2019 | 2020 onwards |
|---|---|---|
| 24-h alive-exit Brier | 0.1249 | 0.1213 |
| 24-h alive-exit C statistic | 0.63 | 0.642 |
| 24-h alive-exit calibration slope | 0.94 | 1.03 |
| 24-h alive exit, predicted − observed | 0.057 | 0.061 |
| 48-h alive-exit Brier | 0.2141 | 0.2072 |
| 72-h alive-exit Brier | 0.2447 | 0.238 |
| 24-h ICU-death Brier / C statistic | 0.0193 / 0.646 | 0.0135 / 0.637 |
| Simulator A − baseline 0, 24-h alive-exit Brier (95% CI) | −0.0011 (−0.0033 to 0.0013) | +0.0048 (+0.0023 to +0.0072) |

Baseline 0 discriminated poorly (C statistic 0.63–0.64, against 0.68–0.75 for the models) but overpredicted exit less than the models (0.057–0.061, against 0.077–0.139), because its rates come from the same landmark and subtype. Its 24-hour Brier score was close to that of the simulator in 2017–2019 and lower than that of every model from 2020 (models 0.1245–0.1390; Table 4). The 24-hour Brier score was therefore dominated by the overall level of exit, which all models overpredicted, rather than by the ranking of patients (see also the post hoc calculation in S4).

**Expected and observed exits.** The plan specified daily expected and observed numbers of exits per unit. Daily unit census cannot be reconstructed in MIMIC-IV, because dates are shifted separately for each patient. The closest feasible quantity is shown in Table S7c.

**Table S7c. Patients in the ICU at each landmark, with expected (sum of simulator A probabilities) and observed numbers leaving alive within 24 hours, temporal validation 2017–2019.**

| Landmark | Unit type | Patients | Expected exits | Observed exits |
|---|---|---|---|---|
| 0 h | Standard ICU | 1,090 | 139 | 60 |
| 0 h | Intermediate / step-down | 624 | 130 | 46 |
| 24 h | Standard ICU | 934 | 180 | 170 |
| 24 h | Intermediate / step-down | 645 | 220 | 131 |
| 48 h | Standard ICU | 647 | 122 | 72 |
| 48 h | Intermediate / step-down | 595 | 202 | 134 |
| 72 h | Standard ICU | 476 | 75 | 45 |
| 72 h | Intermediate / step-down | 532 | 164 | 150 |

#### S8. Ablations

Each reduced simulator was refitted with the changed modules re-selected by the same rule (Table S8b) and evaluated on the same 5,543 temporal-validation patient-landmarks with the same random seeds as the full simulator (paired patient-level bootstrap, 1,000 samples). No reduced simulator produced a logical violation.

**Table S8a. Full and reduced simulators, temporal validation 2017–2019.**

| Measure | Full simulator | A: no readiness inputs in C1 and P1 | B: no clock, weekend or unit in C1 | C: no loss of readiness after confirmation |
|---|---|---|---|---|
| 24-h alive-exit Brier | 0.1238 | 0.1256 | 0.1285 | 0.1261 |
| Difference from full (95% | — | +0.0018 (+0.0002 | +0.0047 | +0.0023 (+0.0016 to |

| Measure | Full simulator | A: no readiness inputs in C1 and P1 | B: no clock, weekend or unit in C1 | C: no loss of readiness after confirmation |
|---|---|---|---|---|
| CI) | | to +0.0034) | (+0.0038 to +0.0055) | +0.0029) |
| 24-h calibration slope | 0.95 | 1.15 | 0.86 | 0.94 |
| 24-h predicted − observed | 0.077 | 0.077 | 0.092 | 0.084 |
| 48-h alive-exit Brier | 0.203 | 0.2019 | 0.2086 | 0.2064 |
| 48-h calibration slope | 0.89 | 1.24 | 0.82 | 0.87 |
| 72-h alive-exit Brier | 0.2234 | 0.2246 | 0.2268 | 0.2274 |
| 72-h calibration slope | 1.01 | 1.43 | 0.94 | 1.0 |
| 24-h C statistic | 0.708 | 0.668 | 0.702 | 0.695 |
| 72-h ICU-death Brier | 0.0429 | 0.0429 | 0.0429 | 0.0429 |
| Length-of-stay Wasserstein distance (h) | 30.4 | 31.6 | 34.9 | 35.9 |
| Median length of stay, simulated / observed (h) | 65 / 90 | 66 / 90 | 61 / 90 | 62 / 90 |
| Readiness relapse, simulated / observed | 0.411 / 0.391 | 0.490 / 0.391 | 0.401 / 0.391 | 0.000 / 0.391 |

Positive differences mean the reduced simulator is worse. Relapse: share of stays simulated from admission that lost readiness for ≥6 hours after confirmation, among those ever ready (in A readiness affects no module and is only recorded; in C it is 0 by design).

**Table S8b. Re-selection of the modules changed in each reduced simulator.**

| Reduced simulator | Module | Selected | Reason |
|---|---|---|---|
| A | C1 | L1 | none eligible, L1 by rule |
| A | P1a | L2 (α=0.0, λ=1.00e+01) | lowest eligible |
| A | P1b | L3 (leaves 31, min samples 1000, 42 trees) | lowest eligible |
| B | C1 | L1 | none eligible, L1 by rule |
| C | D3a | L3 (leaves 7, min samples 1000, 302 trees) | lowest eligible |
| C | D3b | L1 | lowest eligible |
| C | C1 | L1 | none eligible, L1 by rule |
| C | P1a | L1 | none eligible, L1 by rule |

#### S9. Parameter uncertainty

The development set was resampled by patient 200 times and every module refitted with fixed hyperparameters. For 300 temporal-validation patient-landmarks (stratified sample), each refit was simulated 1,000 times.

**Table S9. Width of the 95% parameter-uncertainty interval (97.5th minus 2.5th percentile, probability scale).**

| Prediction | Final-model prediction, median | Interval width, median | Interval width, 90th percentile | Final prediction inside the interval |
|---|---|---|---|---|
| 24-h alive exit | 0.241 | 0.065 | 0.101 | 0.99 |
| 48-h alive exit | 0.454 | 0.083 | 0.122 | 0.997 |
| 72-h alive exit | 0.589 | 0.089 | 0.122 | 0.993 |
| 24-h ICU death | 0.007 | 0.012 | 0.041 | 0.99 |
| 48-h ICU death | 0.019 | 0.021 | 0.064 | 0.993 |
| 72-h ICU death | 0.028 | 0.029 | 0.075 | 0.983 |

These intervals reflect uncertainty from estimating the modules on finite data. They are distinct from the 80% prediction intervals, which describe the spread of an individual's future path.

---

#### S10. Sensitivity analyses

Three sensitivity analyses were pre-specified in the simulator analysis plan. Analyses 1 and 2 repeated the full pipeline (training tables, data audit, module selection, system checks, direct models, final fit and temporal validation) in separate folders; the main analysis was not touched. In both, the strongest direct model on internal validation was the GRU rather than GBM, and the primary comparison was made against it, as the plan requires; the comparison against GBM is also shown.

**Sensitivity analysis 1: split by anchor year group.** Development used the 2008–2010, 2011–2013 and 2014–2016 groups (internal validation = 2014–2016; 3,816 stays); temporal validation used the 2017–2019 group (1,398 stays, 4,546 patient-landmarks). Module selection changed in places (for example, L3 for D1 and C1, and L1 for D2 from states 1 and 3), as expected when the rule is applied to different data.

**Sensitivity analysis 2: physiological readiness in place of stroke-extended readiness.** Readiness in D3 and in every module that uses it was based on the four physiological conditions only. The population and split were those of the main analysis. Ten of the 11 modules kept the same algorithm; P1a changed from L1 to L2.

**Table S10a. Primary comparison in the sensitivity analyses (24-h alive-exit Brier score, four landmarks pooled; 95% CI from 1,000 patient-level bootstrap samples).**

| Analysis | Simulator A | Strongest direct model (GRU) | Difference vs GRU | GBM | Difference vs GBM |
|---|---|---|---|---|---|
| Main analysis | 0.1238 | — (GBM strongest) | — | 0.1245 | −0.0007 (−0.0030 to 0.0016) |
| 1. Anchor-year-group split | 0.1146 | 0.1147 | −0.0000 (−0.0015 to 0.0015) | 0.1135 | +0.0012 (−0.0005 to 0.0029) |
| 2. Physiological readiness | 0.1240 | 0.1253 | −0.0013 (−0.0034 to 0.0009) | 0.1253 | −0.0012 (−0.0032 to 0.0007) |

**Table S10b. Secondary results across analyses (temporal validation).**

| Measure | Main analysis | 1. Anchor-year-group split | 2. Physiological readiness |
|---|---|---|---|
| 48-h alive-exit Brier, simulator A / GBM | 0.203 / 0.2073 | 0.1948 / 0.1908 | 0.2029 / 0.2094 |
| 72-h alive-exit Brier, simulator A / GBM | 0.2234 / 0.2276 | 0.2218 / 0.2162 | 0.2246 / 0.2287 |
| 48-h calibration slope, simulator A / direct models | 0.89 / 0.75–0.84 | 1.0 / 0.75–0.86 | 0.89 / 0.72–0.82 |
| 72-h calibration slope, simulator A / direct models | 1.01 / 0.69–0.86 | 1.11 / 0.69–0.87 | 0.99 / 0.67–0.83 |
| 24-h alive-exit C statistic, simulator A / GBM / GRU | 0.708 / 0.740 / 0.749 | 0.722 / 0.739 / 0.735 | 0.709 / 0.734 / 0.741 |
| 24-h alive exit, predicted − observed (simulator A) | 0.077 | 0.051 | 0.078 |
| Length-of-stay Wasserstein distance, A / B (h) | 30.4 / 38.4 | 29.1 / 30.9 | 34.2 / 42.1 |

The primary comparison remained indistinguishable in both analyses. The simulator kept the calibration slopes closest to 1 at 48 and 72 hours, the direct models kept the higher C statistics, and all models still overpredicted exit. The lowest 48- and 72-hour Brier scores, held by the simulator in the main analysis, moved to GBM under the anchor-year-group split but stayed with the simulator when physiological readiness was used.

**Sensitivity analysis 3: 100 instead of 200 bootstrap samples for parameter uncertainty.** The first 100 of the 200 resamples were used to recompute the 95% intervals for the same 300 patient-landmarks.

**Table S10c. Parameter-uncertainty intervals with 200 and 100 bootstrap samples.**

| Prediction | Median width, 200 | Median width, 100 | Relative difference in width (median) | Absolute difference in end points (median) | Final prediction inside the interval, 200 / 100 |
|---|---|---|---|---|---|
| 24-h alive exit | 0.065 | 0.063 | 5.0% | 0.0015 | 99.0% / 98.7% |
| 48-h alive exit | 0.083 | 0.083 | 4.6% | 0.0021 | 99.7% / 99.0% |
| 72-h alive exit | 0.089 | 0.089 | 4.2% | 0.0022 | 99.3% / 99.3% |
| 24-h ICU death | 0.012 | 0.012 | 4.8% | 0.0005 | 99.0% / 98.0% |
| 48-h ICU death | 0.021 | 0.021 | 4.2% | 0.0005 | 99.3% / 99.0% |
| 72-h ICU death | 0.029 | 0.029 | 4.8% | 0.0010 | 98.3% / 98.3% |

#### S11. Results for 2020 onwards

Stays with approximate admission year 2020 or later (1,521 stays, 4,979 patient-landmarks), evaluated with the models frozen on the development set. Primary difference, simulator A − GBM: +0.0002 (95% CI −0.0026 to 0.0027).

**Table S11a. Length of stay (h): observed and simulated percentiles.**

| Percentile | Observed | Simulator A | Simulator B (all L1) |
|---|---|---|---|
| 10th | 28 | 17 | 18 |
| 25th | 47 | 32 | 31 |
| 50th | 95 | 65 | 63 |
| 75th | 194 | 134 | 120 |
| 90th | 360 | 260 | 230 |

Wasserstein distance over the full stay: simulator A 41.6 h, B 49.6 h. Up to 72 h: A 7.92, B 8.26, LR 9.45, GBM 7.40.

**Table S11b. 24-h alive-exit Brier score by subgroup.**

| Subgroup | Patient-landmarks | Simulator A | Simulator B | LR | GBM | GRU |
|---|---|---|---|---|---|---|
| Standard ICU | 2,797 | 0.0983 | 0.0985 | 0.0951 | 0.0978 | 0.1089 |
| Intermediate / step-down | 2,182 | 0.1618 | 0.1629 | 0.1622 | 0.1621 | 0.1776 |
| AIS | 2,764 | 0.1433 | 0.1443 | 0.1431 | 0.1443 | 0.1551 |
| ICH | 1,597 | 0.122 | 0.1222 | 0.1194 | 0.1215 | 0.1392 |
| SAH | 618 | 0.06 | 0.0599 | 0.0543 | 0.0552 | 0.067 |
| Landmark 0 h | 1,521 | 0.07 | 0.0683 | 0.0731 | 0.0598 | 0.0621 |
| Landmark 24 h | 1,416 | 0.1629 | 0.1642 | 0.1627 | 0.1613 | 0.1796 |
| Landmark 48 h | 1,124 | 0.1449 | 0.1473 | 0.1389 | 0.1543 | 0.1706 |
| Landmark 72 h | 918 | 0.1393 | 0.1406 | 0.1329 | 0.1463 | 0.1653 |

**Table S11c. Remaining ICU time (simulator A).**

| Way the stay ended | Patient-landmarks | Mean absolute error of the predicted median (h) | 80% prediction-interval coverage |
|---|---|---|---|
| Alive exit | 4,532 | 96.8 | 0.786 |
| ICU death | 387 | 93.9 | 0.739 |

For comparison, the same subgroup results for the main temporal validation (2017–2019) are: standard ICU 0.0923 (simulator A) vs 0.0907 (GBM); intermediate 0.1652 vs 0.1689; AIS 0.1378 vs 0.1411; ICH 0.1177 vs 0.1141; SAH 0.065 vs 0.0628. Remaining-time coverage 2017–2019: alive exits 0.813, ICU deaths 0.749.

---

#### S12. Post-exit modules

P1a predicts direct hospital discharge within 1 hour of ICU exit; P1b predicts, hour by hour for 72 hours after exit, ICU readmission in the same hospitalisation, in-hospital death or discharge. Both use the patient's situation at exit: condition state, readiness at exit and length of stay, and, for P1b, the hour and unit type of exit.

**Table S12. Post-exit predictions of simulator A among alive ICU exits.**

| Outcome | 2017–2019: predicted / observed | 2017–2019: C statistic | 2020 onwards: predicted / observed | 2020 onwards: C statistic |
|---|---|---|---|---|
| Direct hospital discharge (P1a) | 0.059 / 0.315 | 0.544 | 0.06 / 0.258 | 0.483 |
| ICU readmission within 72 h (P1b) | 0.058 / 0.075 | 0.629 | 0.058 / 0.068 | 0.613 |
| In-hospital death within 72 h (P1b) | 0.038 / 0.041 | 0.935 | 0.031 / 0.043 | 0.927 |

The direct-discharge module was fitted on years in which direct discharge from an ICU-level unit was rare (5.9% of alive exits in the development set; Table 1) and failed in both validation sets.

---

#### S13. Odds ratios, effect plots and SHAP

Odds ratios come from the final modules fitted on the whole development set. State codes are as in S5 (0 renal dysfunction, 1 respiratory support, 2 impairment with low support, 3 preserved). L1 modules are unpenalised logistic regressions (point estimates); multinomial modules are shown relative to their first outcome class. L2 coefficients are penalised (ridge, λ=10) and are shown on the odds-ratio scale only to indicate direction. Spline terms (hours since ICU admission) are shown in Figure S1. Modules selected as LightGBM (D2 from state 1, D3a, D3b, P1b) are summarised with TreeSHAP in Table S13h and Figure S2.

**Table S13a. D1, death in the ICU (L1): odds ratios.**

| Term | Death |
|---|---|
| Condition state 0 (vs before hour 6) | 1.96 |
| Condition state 1 (vs before hour 6) | 3.70 |
| Condition state 2 (vs before hour 6) | 0.36 |
| Condition state 3 (vs before hour 6) | 0.03 |
| ICH (vs AIS) | 1.46 |
| SAH (vs AIS) | 1.49 |
| Neuro intermediate / step-down unit | 0.78 |
| Age, per 10 years | 1.33 |

**Table S13b. D2-0, condition state at hour 6 (L1): odds ratios.**

| Term | State 1 vs 0 | State 2 vs 0 | State 3 vs 0 |
|---|---|---|---|
| ICH (vs AIS) | 1.96 | 1.22 | 1.59 |
| SAH (vs AIS) | 2.50 | 1.61 | 2.40 |
| Neuro intermediate / step-down unit | 0.38 | 3.42 | 4.61 |
| Reperfusion therapy | 1.37 | 2.16 | 2.84 |
| Admitted from operating room (vs emergency department) | 0.99 | 0.79 | 1.09 |
| Admitted from direct admission (vs emergency department) | 0.35 | 0.53 | 0.48 |
| Admitted from earlier ICU stay (vs emergency department) | 1.95e-03 | 0.02 | 0.19 |
| Admitted from ward (vs emergency department) | 0.81 | 0.96 | 1.16 |
| Age, per 10 years | 1.00 | 1.33 | 0.93 |

**Table S13c. D2 from state 0, renal dysfunction (L2, penalised): odds ratios.**

| Term | To 1 vs 0 | To 2 vs 0 | To 3 vs 0 |
|---|---|---|---|
| ICH (vs AIS) | 1.00 | 1.00 | 1.00 |
| SAH (vs AIS) | 1.01 | 1.01 | 1.01 |
| Neuro intermediate / step-down unit | 1.00 | 1.01 | 1.02 |
| Age, per 10 years | 1.00 | 1.00 | 1.00 |

**Table S13d. D2 from state 2, impairment, low support (L1): odds ratios.**

| Term | To 1 vs 0 | To 2 vs 0 | To 3 vs 0 |
|---|---|---|---|
| ICH (vs AIS) | 1.37 | 1.27 | 1.20 |
| SAH (vs AIS) | 2.52 | 1.71 | 1.57 |
| Neuro intermediate / step-down unit | 1.00 | 5.71 | 3.27 |
| Age, per 10 years | 1.11 | 1.16 | 0.99 |

**Table S13e. D2 from state 3, preserved (L2, penalised): odds ratios.**

| Term | To 1 vs 0 | To 2 vs 0 | To 3 vs 0 |
|---|---|---|---|
| ICH (vs AIS) | 1.00 | 1.00 | 1.00 |
| SAH (vs AIS) | 1.00 | 1.00 | 1.00 |
| Neuro intermediate / step-down unit | 1.00 | 1.00 | 1.01 |
| Age, per 10 years | 1.00 | 1.00 | 1.00 |

**Table S13f. C1, destination (L1): odds ratios.**

| Term | Move vs stay | Leave vs stay |
|---|---|---|
| Readiness ever confirmed | 1.66 | 2.17 |
| 0–6 h since first confirmation (vs never confirmed) | 0.99 | 0.87 |
| 6–24 h since first confirmation (vs never confirmed) | 1.36 | 1.51 |
| 24–72 h since first confirmation (vs never confirmed) | 1.28 | 1.41 |
| ≥72 h since first confirmation (vs never confirmed) | 0.97 | 1.17 |
| Readiness met now | 2.74 | 3.02 |
| Condition state 0 (vs before hour 6) | 0.45 | 0.74 |
| Condition state 1 (vs before hour 6) | 0.34 | 0.69 |
| Condition state 2 (vs before hour 6) | 0.93 | 1.10 |
| Condition state 3 (vs before hour 6) | 1.01 | 1.66 |
| Hour 04:00–08:00 (vs 00:00–04:00) | 0.21 | 0.08 |
| Hour 08:00–12:00 (vs 00:00–04:00) | 0.94 | 1.33 |
| Hour 12:00–16:00 (vs 00:00–04:00) | 2.03 | 5.47 |
| Hour 16:00–20:00 (vs 00:00–04:00) | 2.49 | 11.61 |

| Term | Move vs stay | Leave vs stay |
|---|---|---|
| Hour 20:00–24:00 (vs 00:00–04:00) | 1.71 | 6.22 |
| Weekend | 0.74 | 0.98 |
| Neuro intermediate / step-down unit | 1.23 | 0.80 |
| ICH (vs AIS) | 0.77 | 0.99 |
| SAH (vs AIS) | 0.45 | 0.40 |

**Table S13g. P1a, direct hospital discharge (L1): odds ratios.**

| Term | Direct discharge |
|---|---|
| Condition state at exit 0 (vs none) | 0.55 |
| Condition state at exit 1 (vs none) | 0.70 |
| Condition state at exit 2 (vs none) | 0.26 |
| Condition state at exit 3 (vs none) | 0.48 |
| Readiness lost at exit (vs met) | 2.69 |
| Readiness never met (vs met) | 3.75 |
| ICH (vs AIS) | 1.14 |
| SAH (vs AIS) | 0.83 |
| log ICU length of stay | 2.12 |
| Age, per 10 years | 1.03 |

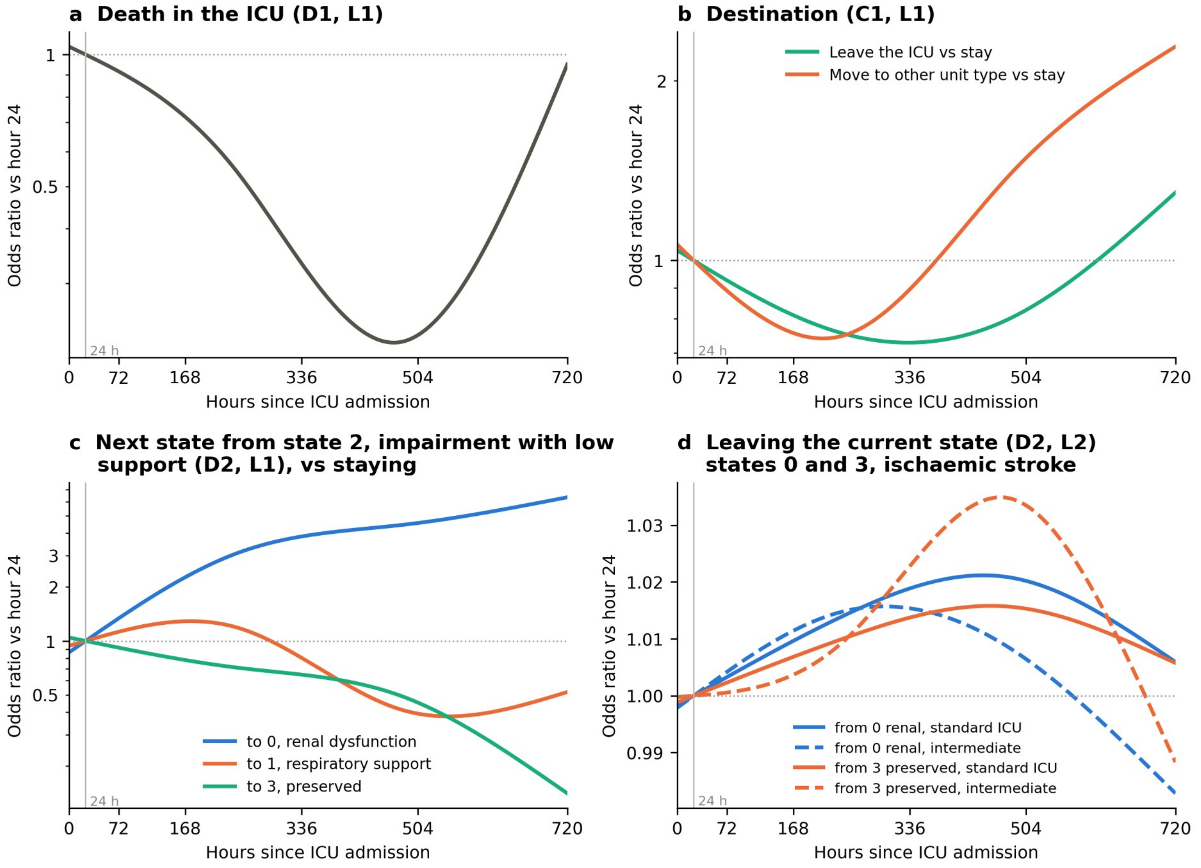

**Figure S1. Effect of hours since ICU admission in the L1 and L2 modules.** Odds ratios relative to hour 24 (log scale, except panel d). (a) Death in the ICU. (b) Destination: leaving the ICU and moving to the other unit type, each against staying. (c) Next condition state from state 2 (impairment with low support), against staying in state 2. (d) Leaving the current state from states 0 (renal dysfunction) and 3 (preserved) (penalised elastic net; the curves are nearly flat and should be read for direction only), standard ICU and intermediate unit, ischaemic stroke. Curves beyond about 14 days rest on few stays. Drawn from the frozen model parameters; no patient data were used.

**Table S13h. LightGBM modules: TreeSHAP importance and agreement with the L1 model.** Mean absolute SHAP value on the log-odds scale (multiclass: average over classes after centring), in a random sample of up to 40,000 development rows. Agreement is the Spearman correlation, row by row, between a variable's SHAP contribution and its contribution in the L1 version of the same module.

| Module | Rank | Variable | Mean \|SHAP\| | Agreement with L1 |
|---|---|---|---|---|
| D2 from state 1 | 1 | Hours in ICU | 0.206 | 0.78 |
| D2 from state 1 | 2 | Age (per 10 y) | 0.090 | 0.46 |
| D2 from state 1 | 3 | Stroke subtype | 0.069 | 0.75 |
| D2 from state 1 | 4 | Intermediate unit | 0.000 | — |
| D3a, readiness gained | 1 | Hours in current readiness run | 0.776 | 0.94 |
| D3a, readiness gained | 2 | Condition state | 0.516 | 0.89 |
| D3a, readiness gained | 3 | Hours in ICU | 0.187 | 0.21 |
| D3a, readiness gained | 4 | Reperfusion, first 24 h | 0.131 | 0.34 |
| D3a, readiness gained | 5 | Stroke subtype | 0.060 | 0.67 |
| D3a, readiness gained | 6 | Intermediate unit | 0.004 | 0.18 |
| D3b, readiness lost | 1 | Hours in current readiness run | 0.584 | 0.98 |
| D3b, readiness lost | 2 | Hours in ICU | 0.150 | 0.90 |
| D3b, readiness lost | 3 | Condition state | 0.146 | 0.73 |
| D3b, readiness lost | 4 | Stroke subtype | 0.133 | 0.92 |
| D3b, readiness lost | 5 | Intermediate unit | 0.083 | 0.47 |
| D3b, readiness lost | 6 | Reperfusion, first 24 h | 0.000 | — |
| P1b, post-exit outcomes | 1 | Hours since ICU exit | 0.360 | 0.55 |
| P1b, post-exit outcomes | 2 | Age (per 10 y) | 0.163 | 0.45 |
| P1b, post-exit outcomes | 3 | Condition state at exit | 0.160 | 0.61 |
| P1b, post-exit outcomes | 4 | Readiness at exit | 0.141 | 0.71 |
| P1b, post-exit outcomes | 5 | log ICU length of stay | 0.089 | -0.08 |
| P1b, post-exit outcomes | 6 | Exit 19:00–07:00 | 0.036 | 0.45 |
| P1b, post-exit outcomes | 7 | Stroke subtype | 0.032 | 0.44 |
| P1b, post-exit outcomes | 8 | Reperfusion therapy | 0.026 | 0.32 |
| P1b, post-exit outcomes | 9 | Exit from intermediate unit | 0.020 | 0.29 |

Every variable acted in the same direction as in the L1 version (Spearman 0.18–0.98) except log ICU length of stay in P1b (−0.08). A dash marks a variable that the model did not use (SHAP 0). SHAP values describe the model, not causal effects.

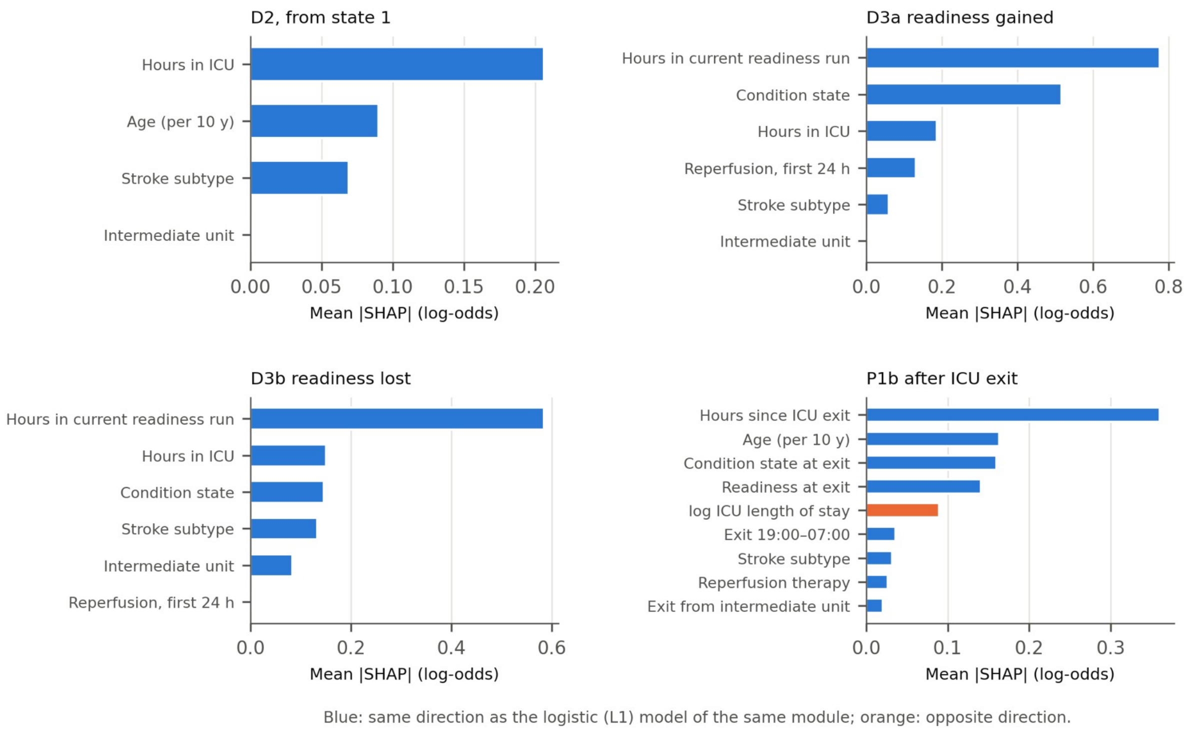

**Figure S2. Mean absolute SHAP values of the LightGBM modules.**

#### S14. Applicability flags

Every prediction is flagged as outside the model's validated range when any of the following holds.

**Table S14. Applicability flags.**

| Flag | Rule |
|---|---|
| Unit not in the list | The current unit is not one of the units in Table S1 |
| Beyond 30 days | The patient has been in the ICU for more than 720 hours |
| Missing inputs | An input is missing beyond the pre-specified rules (for example, no condition state after hour 6) |
| Off-landmark | The prediction time is not one of the four landmarks (0, 24, 48, 72 h); predictions at other hours come from the same engine but were not validated |
| Post-exit beyond 72 h | Outcomes more than 72 hours after exit, and a second ICU stay after readmission, are not simulated |

#### S15. Monitoring intensity in T3 hours

An hour in T3 was classed as high-intensity monitoring when 3 or more GCS assessments had been recorded in the preceding 6 hours (amendment 1.2 of the ICU-time analysis plan). A sensitivity definition used 2 or more assessments in the preceding 4 hours.

**Table S15a. High-intensity monitoring in T3 hours by unit type.**

| Unit type | T3 hours | High intensity, % of hours | High intensity, mean % per patient | Sensitivity definition, % of hours |
|---|---|---|---|---|
| Standard ICU | 208,889 | 59.1 | 54.6 | 62.0 |
| Neuro intermediate / step-down | 206,047 | 49.2 | 50.3 | 52.1 |
| All | 414,962 | 54.2 | 51.3 | 57.1 |

**Table S15b. High-intensity monitoring in T3 hours by day of stay (% of hours).**

| Unit type | Day 1 | Days 2–3 | Days 4–7 | Days 8–14 | Days 15–30 |
|---|---|---|---|---|---|
| Standard ICU | 69.7 | 68.7 | 59.7 | 46.2 | 24.1 |
| Neuro intermediate / step-down | 66.0 | 63.5 | 48.9 | 38.5 | 22.2 |

A small number of T3 hours could not be matched to a unit type; their high-intensity share was 90.9% on day 1 and 100.0% on days 2–3. They are included in "All".

---

#### S16. Reporting checklists

Locations refer to the sections of the main text and of this supplement. "Not applicable" and "not addressed" are stated where they apply.

**Table S16a. TRIPOD+AI (Collins et al., BMJ 2024;385:e078378).**

| Item | Topic | Where reported |
|---|---|---|
| 1 | Title | Title (simulation and temporal validation; ICU trajectories after stroke; outcome = ICU exit) |
| 2 | Abstract | Structured abstract |
| 3a | Healthcare context and rationale, existing models | Background and Significance, paragraphs 1–3 (refs 1–5) |
| 3b | Target population, intended purpose and users | Background; Objective; Discussion, Simulator and direct prediction |
| 3c | Known health inequalities | Not addressed: the study did not examine sociodemographic inequalities |
| 4 | Objectives, development and/or validation | Objective |
| 5a | Data sources for development and evaluation | Methods, Data and population; Temporal split |
| 5b | Dates of data | Methods, Temporal split; Table 1; S11 |
| 6a | Setting, number and location of centres | Methods, Data and population (one centre, Boston) |
| 6b | Eligibility | Methods, Data and population; earlier study (ref 10) |
| 6c | Treatments and their handling | Methods, Operational definition of transfer readiness (treatments as readiness criteria); S2 |
| 7 | Data pre-processing and quality checks | Methods; S2; S4 (data audit, label checks); preprocessing of condition states from ref 10 |
| 8a | Outcome and time horizon | Methods, Comparators and evaluation (alive exit and ICU death within 24, 48, 72 h); S3 Part B, B7 |
| 8b | Outcome assessors | Not applicable: outcomes taken from recorded times |
| 8c | Blinding of outcome assessment | Not applicable: outcomes taken from recorded times |
| 9a | Choice of initial predictors | Methods, Simulator structure; Table 2; S3 Part B, B3 |
| 9b | Definition of predictors | Table 2; S2; S5 |
| 9c | Predictor assessors | Not applicable: recorded data |
| 10 | Study size | All eligible stays in the database (Methods, Data and population); no sample-size calculation; events per split in S3 Part B, B2 |
| 11 | Missing data | S2 (validity windows, unknown GCS counted as not met); S3 Part B, B3; S14 |
| 12a | Use of the data, partitioning | Methods, Temporal split; Algorithms, tuning and selection |
| 12b | Handling of predictors | S5 (splines, coding); S3 Part B, B4.2 |
| 12c | Model type, building steps, tuning, internal validation | Methods, Algorithms, tuning and selection; S5; S3 Part B, B4 |

| Item | Topic | Where reported |
|---|---|---|
| 12d | Heterogeneity across clusters | Not applicable: single centre; results by unit type and stroke subtype in S11 |
| 12e | Performance measures and comparison of models | Methods, Comparators and evaluation |
| 12f | Model updating | Not done: frozen model evaluated without updating |
| 12g | How predictions were calculated | Methods, Simulator structure (1,000 simulated paths per patient); S3 Part B, B5 |
| 13 | Class imbalance | No class-imbalance methods (S3 Part B, B4.2: no class weights) |
| 14 | Fairness | Not addressed |
| 15 | Model output | Methods, Simulator structure (probabilities and remaining-time distribution) |
| 16 | Differences between development and evaluation data | Results, Population; Table 1; Discussion, Detecting changes in care structure |
| 17 | Ethics approval and consent | Ethics approval |
| 18a | Funding | Funding |
| 18b | Conflicts of interest | Conflict of interest |
| 18c | Protocol | S3 (statistical analysis plans) |
| 18d | Registration | Ethics approval (not registered) |
| 18e | Data sharing | Data availability |
| 18f | Code sharing | Data availability |
| 19 | Patient and public involvement | Patient and public involvement (none) |
| 20a | Flow of participants | Methods, Data and population; Figure 1a |
| 20b | Characteristics, events, follow-up | Table 1 |
| 20c | Comparison of development and evaluation data | Table 1 (by split) |
| 21 | Numbers of participants and events in each analysis | Methods, Temporal split; Table 4; S3 Part B, B2; S5–S7 |
| 22 | Full model specification | S5, S13; frozen model package (Data availability) |
| 23a | Performance with confidence intervals, subgroups | Results, Temporal validation; Table 4; Figure 4; S8, S10, S11 (subgroup estimates without confidence intervals) |
| 23b | Heterogeneity across clusters | Not applicable |
| 24 | Model updating results | Not applicable |
| 25 | Interpretation | Discussion |
| 26 | Limitations | Discussion, Limitations |
| 27a | Poor-quality or unavailable input data in use | S14 (applicability flags); S2 (missing-data rules) |
| 27b | User interaction and expertise | Not applicable: research model, not deployed |
| 27c | Next steps | Discussion, Detecting changes in care structure (external validation in eICU); Limitations |

**Table S16b. STRESS-DES (Monks et al., J Simul 2019;13:55–67). The simulator is an individual-level, discrete-time simulation; items on resources and queues do not apply.**

| Item | Topic | Where reported |
|---|---|---|
| 1.1 | Purpose of the model | Objective; Background |
| 1.2 | Model outputs | Methods, Simulator structure; Comparators and evaluation |
| 1.3 | Experimentation aims | Methods (ablations and sensitivity analyses); S3 Part B, B6.3, B10 |
| 2.1 | Base model overview diagram | Figure 1b |
| 2.2 | Base model logic | Methods, Simulator structure; S3 Part B, B2.1 and B5 |
| 2.3 | Scenario logic | Methods (ablations A–C); S8 |
| 2.4 | Algorithms | S5; S3 Part B, B3–B4 |

| Item | Topic | Where reported |
|---|---|---|
| 2.5.1 | Entities | Patients (ICU stays) with the attributes in Table 2 |
| 2.5.2 | Activities | Hourly modules D1, D2-0/D2, C1, D3a/D3b, P1a/P1b (Table 2; Figure 1b) |
| 2.5.3 | Resources | Not modelled (no bed or staff capacity) |
| 2.5.4 | Queues | Not modelled |
| 2.5.5 | Entry and exit points | Entry at ICU admission or a landmark; exit by alive ICU exit, ICU death or truncation at 720 h; post-exit phase up to 72 h (S3 Part B, B2.1) |
| 3.1 | Data sources | Methods, Data and population; Temporal split |
| 3.2 | Pre-processing | S2; S4; earlier study (ref 10) |
| 3.3 | Input parameters | S5, S13 |
| 3.4 | Assumptions | S2 (readiness rules); S3 Part B, B3.2 (state transitions conditional on remaining in the ICU); Limitations |
| 4.1 | Initialisation | Terminating system; no warm-up; each path starts from the patient's observed state at the landmark (S3 Part B, B5) |
| 4.2 | Run length | Up to 720 hours in the ICU plus 72 hours after exit; hourly time step |
| 4.3 | Estimation approach | 1,000 simulated paths per patient-landmark; parameter uncertainty from 200 bootstrap refits (S9) |
| 5.1 | Software | Methods, Software (Python 3.9, pandas, scikit-learn 1.6.1, LightGBM 4.6.0, PyTorch 2.8.0) |
| 5.2 | Random sampling | NumPy default generator (PCG64); one seed per patient-landmark (stay identifier × 100 + landmark), identical across compared simulators (S8) |
| 5.3 | Model execution | Fixed six-step order within each hour (S3 Part B, B2.1); logic checks (S6; S3 Part B, B7.3) |
| 5.4 | System specification | Local workstation, 18 cores, 48 GB memory (S3 Part B, B12); run times in S4 |
| 6.1 | Model sharing | Data availability (code and frozen model package) |

**Table S16c. RECORD items additional to STROBE (Benchimol et al., PLoS Med 2015;12:e1001885).**

| Item | Topic | Where reported |
|---|---|---|
| 1.1 | Type of data in title or abstract | Abstract, Materials and Methods (MIMIC-IV) |
| 1.2 | Geographic region and timeframe | Methods, Data and population (Boston, 2008–2022) |
| 1.3 | Linkage between databases | MIMIC-IV and MIMIC-IV-ED, linked within PhysioNet (Methods, Data and population) |
| 6.1 | Methods of population selection | Methods, Data and population; codes in the earlier study (ref 10) |
| 6.2 | Validation of codes | Not done in this study; see the earlier study (ref 10) |
| 6.3 | Linkage flow diagram | Not applicable (linkage within one database family) |
| 7.1 | Codes and algorithms | Earlier study (ref 10) for diagnosis codes; S1 (units); S2 (readiness items) |
| 12.1 | Investigators' access to the database | Full access to the credentialed MIMIC-IV release (Ethics approval) |
| 12.2 | Data cleaning | S2; S4; earlier study (ref 10) |
| 12.3 | Linkage methods | Database identifiers (subject, admission and stay identifiers) |
| 13.1 | Selection of persons, flow | Methods, Data and population; Figure 1a |
| 19.1 | Implications of using routine data | Discussion, Limitations (what the readiness definition can see; approximate admission years) |
| 22.1 | Access to protocol, data and code | S3; Data availability |
